# How COVID-19 vaccination in England varied by geography, demography, deprivation and registry: Bayesian ecological modelling

**DOI:** 10.64898/2025.12.01.25341337

**Authors:** Greg Dropkin

## Abstract

**Background:** COVID-19 vaccines were freely available in England through the NHS with initial rollout from late 2020, but uptake was highly variable by local geography. Do census variables and deprivation explain the variation in uptake?

**Methods:** Middle Super Output Area level data for 1st and 2nd Primaries (P1, P2) and Booster / 3rd Injection (B3I) was obtained from the COVID-19 Dashboard. Covariate sources included Census 2021, the Index of Multiple Deprivation (IMD), flu vaccination, the EU Referendum, military bases and prisons, and the ratio of the vaccine registry to the Census population aged 12+ which was designated VRx. A Bayesian multilevel beta-binomial model including factors for lower tier local authority nested within region was fitted to the uptake. Leave-one-out methods were used to measure the impact of model terms and groups and identify possible outliers.

**Results:** The model passed Bayesian tests for convergence and fit the data well with few outliers. P1 uptake was the key predictor of P2, which predicted B3I. Regional and Local Authority factors and interaction of IMD with VRx had strong impacts on P1. The area proportions of various ethnicities, younger age, international migration, voting to “Leave” the EU, and declining census questions on religion or sexual orientation, were all negative predictors of P1 uptake. Amongst ethnicities the “Other White” group had the strongest impact. The “random effects” for London and the South West were credibly higher than those for North West or North East. Differences between the ten highest and lowest nested local authority effects were credible and generally larger than regional differences.

**Conclusions:** Modelling can adjust for uncertainty in Vaccine Register data. Vaccination pandemic planning should prioritise the initial phase. Distrust is a credible predictor. Ethnic minorities and in particular “Other White” communities and international migrants need active outreach to overcome barriers including language and the Hostile Environment. Local authority effects and outliers may reflect differences in NHS and public health strategy and capacity to offer relevant provision during the most critical period of vaccination, its outset.

## 1 Background

Vaccines have been a key part of the response to the global pandemic of Coronavirus Disease 2019 (COVID-19) caused by the virus SARS-CoV-2. In many parts of the world, uptake was restricted by insufficient supplies or cost to the patient. In England everyone had free access to prevention and treatments for the disease through the NHS. However, when vaccines were deployed patients faced obstacles including digital exclusion, language barriers, travel costs, time pressures, the Hostile Environment, fear of the vaccines, the disease and contact with the NHS.

In England, the 1st Primary was rolled out from late 2020 and the 2nd Primary followed soon after. The Booster / 3rd Injection was available from Autumn 2021.[1, p26] Before rollout began, the UK Scientific Advisory Group for Emergencies (SAGE) drew on attitude surveys and recognised that certain groups would be less likely to accept the vaccine. In some cases this overlapped with higher levels of disease and mortality.[2, 3]

During the vaccination programme, researchers explored how ethnicity and migration affected uptake, sometimes controlling for other demographic and socio-economic factors.[4–8] Uptake also shows striking geographic variation, even between adjacent neighbourhoods. Is this fully explained by demography and deprivation? Which aspects of demography are most relevant? Did the patterns of variation change over time? The Vaccine Registry, which forms the denominator for the uptake, can have errors. Can we adjust for this uncertainty?

Such questions can be addressed using public data at Middle Super Output Area (MSOA) level, a unit of English census geography with around 8000 persons on average. COVID-19 vaccine uptake and a wide range of Census 2021 covariates and deprivation indices are publicly available at this level.

Regions and local authorities within them also show unexplained variation in uptake.[9–12] A multilevel (mixed) model can describe the distribution of such “random effects” and estimate them for a particular region or locality. A multilevel model using splines to incorporate non-linear effects for selected covariates can be specified and fitted as a Generalized Additive Model (GAM).[13] Implementing this in a Bayesian framework [14] improves its accuracy.

As with any observational analysis of grouped data, an association of an outcome with some covariate may be due to its correlation with other covariates which actually cause the effect. I refer to a covariate’s “impact” rather than its “effect”.

## 2 Methods

MSOA-level data on vaccination and the Vaccine Registry was published on the UK government COVID-19 Dashboard during the pandemic. Data from 13 December 2023 was obtained on 14 December 2023. The archive of such data is now the UK Health Security Agency data dashboard.[15]

The downloaded Dashboard data covers the cumulative values of the 1st and 2nd Primary (P1 and P2) and the Booster / 3rd Injection (B3I) and the Vaccine Registry (VR) at 13 December 2023. The rows for Isles of Scilly and City of London were excluded from the analysis, leaving 6789 MSOA. The exclusions were for compatibility with a previous paper [10] but involve only 2053 + 8583 = 10636 persons and are unlikely to influence results here. The Index of Multiple Deprivation (IMD) published in 2019 [16] covers Lower Super Output Areas (LSOA). Census 2021 [17] was conducted on 21 March 2021 and includes data for 6856 MSOA, downloaded via Nomis.[18] The difference in geography is due to boundary changes as some MSOAs were split, merged, or renamed. Local authority data on the European Union (EU) Referendum (2016) was downloaded from the Electoral Reform Commission [19]. Data on influenza vaccination uptake in persons over 65 was available at Unitary Authority level from Public Health England [20]. Data on proximity to military sites was compiled using Census data on Communal Defence Establishments, Census maps and public information from the Services.

The initial dataset contains 6789 rows, named to identify each MSOA by its unique 8-digit geographic code of the form E0200xxxx. The first four columns contain P1, P2, B3I and VR from the Dashboard. Subsequent columns contain numerical covariates and factors.

Deprivation data was obtained for each MSOA by taking a population-weighted average over those LSOA within the MSOA. To obtain census variables, 114 MSOA codes which do not appear in the Census data were identified. In 6 cases a new code was assigned to the old MSOA. In 76 cases an old MSOA was split into two or more new MSOA. Census data for the new MSOAs was summed and, if appropriate, divided by the combined population. In 16 pairs, two old MSOAs were merged and assigned to one new MSOA, whose census variables were assumed to pertain to each of the old MSOAs.

The mergers reduced the dataset from 6789 to 6773 rows. Any non-census variables obtained for the 6789 were combined for the mergers with weighting by VR. Lower Tier Local Authority is treated as a factor **LTLA** with 307 levels, and Region is a factor **REG** with 9 levels. Each MSOA belongs to one **LTLA** within one **REG**. The reduced dataset *dat1* with 6773 rows is indexed by the same geographic codes as in the 6789 row version except for the 16 mergers which are coded as per their new MSOA as shown in the Census. This dataset is Additional file 1; its column names and brief descriptions are shown in Additional file 2 and a table of levels for **LTLA** nested within **REG** is in Additional file 3. This paper uses only some of the covariates in *dat1*.

### Statistical analysis

Preliminary analysis specified and fitted GAMs using the R programme mgcv (Mixed GAM Computation Vehicle).[21] The response consists of integer pairs (P1,VR-P1) for each MSOA, suggesting a binomial model which could take the form:

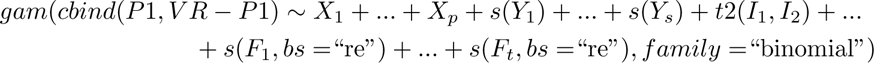

Here gam is the mgcv programme for fitting GAMs. The response term before the ∼ is a matrix with N rows and two columns, where N is the number of MSOA being modelled. *X*_1_ … *X_p_* are parametric terms which contribute to the linear predictor via a single unknown parameter (their slope), *s*(*Y*_1_) … *s*(*Y_s_*) are smooth terms whose contribution is an unknown linear combination of splines approximating a smooth curve, *t*2(*I*_1_*, I*_2_)… are interaction terms composed of a tensor product of splines, and *s*(*F*_1_*, bs* =“re”)… are “random effects” for factors. Fitting the model estimates the unknown parameters, including smoothing parameters governing all the non-parametric terms and penalising their departure from linearity. The resulting terms incorporating the parameter estimates are summed to form the linear predictor *u*, which is then transformed by an inverse link function *inv logit*(*u*) = (1 + *exp*(−*u*))*^−^*^1^ to give the estimated probability of success in the binomial distribution. Models of this type were explored to identify key covariates within *dat1* and to choose appropriate basis dimensions for the smooth terms.

This approach has several problems. The binomial family can underestimate the variance in the data. The more appropriate “beta-binomial” family allows the probability of success to vary by drawing *π* ∼ *B*(*α, β*) from a beta distribution before applying the probability *π* in the binomial step. Here *α* and *β* will depend on the covariates. The beta-binomial family is not available in mgcv. The ratio P1/VR can be modelled as beta-distributed, but this loses the information carried by VR. **LTLA** is nested within **REG**, but modelling with nested factors requires a related programme `gamm4` (Generalized Additive Mixed Models using ‘mgcv’ and ‘lme4’) which interprets all models as multilevel. It is considerably slower than gam.

Both gam and gamm4 are based on choosing a criterion used to optimise the smoothing parameters and all other parameters as a consequence. The fitted model describes the behaviour around the optimised parameter values. To obtain approximate credible intervals, the techniques invoke the asymptotic behaviour as *N* → ∞ and the parameters tend to a normal distribution. But how do we know if our data has enough records to justify the assumption? Despite these problems, mgcv is an excellent fast starting point when considering which covariates to include and how to treat them in the model.

The Bayesian programme `brms` (Bayesian Regression Modelling using ‘Stan’)[22] allows multilevel models with nested factors along with parametric and smooth terms and their interactions. The matrix of response pairs (*P* 1*_i_, V R_i_* − *P* 1*_i_*) can be modelled with a beta-binomial family parametrized by *ϕ* = *α_i_* + *β_i_* and *µ_i_* = *α_i_/ϕ* so that *α_i_* = *µ_i_*∗*ϕ* and *β_i_* = (1−*µ_i_*)∗*ϕ*. Whilst *ϕ* is a single global parameter whose distribution is obtained when fitting the model, *µ_i_* depends on the covariates and its distribution will vary across the MSOA. The corresponding log-likelihood is *lchoose*(*V R_i_, P* 1*_i_*) + *lbeta*(*P* 1*_i_* + *α_i_, V R_i_* − *P* 1*_i_* + *β_i_*) − *lbeta*(*α_i_, β_i_*) where *lchoose*(*n, k*) = *log*(Γ(*n* + 1)) − *log*(Γ(*k* + 1)) − *log*(Γ(*n* − *k* + 1)) and *lbeta*(*a, b*) = *log*(Γ(*a*)) + *log*(Γ(*b*)) − *log*(Γ(*a* + *b*)). The distribution of the modelled uptake at MSOA_i_ is the distribution of *inv logit*(*µ_i_*).

Given the model and data, `brms` generates code to obtain Markov Chain Monte Carlo samples from the posterior distribution of all parameters, using Hamiltonian Monte Carlo implemented in Stan (Software for Bayesian Data Analysis) rather than Metropolis-Hastings or Gibbs sampling. This avoids problems of highly autocorrelated samples which explore the posterior distribution only slowly. For details of the theory and implementation see [14, 23, 24]. Although `brms` / `Stan` is more flexible and accu-rate than `mgcv`, it is much slower as samples are obtained individually rather than by a single fit to the entire data.

The full specification for the main `brms` model ℳ1 and all codes required for the corresponding results are at Additional file 4 with explanatory comments. The model uses parametric terms concerning the “Leave” vote in the 2016 EU Referendum, selected minor occupational groups, communal defence establishments and prisons, indicators for proximity to military bases of the 4 services, Long Term Conditions, uptake of Flu Vaccine, internal UK migration, no passport; smooth univariate terms for Female, three age groups, international migration, No Answer to census questions on Religion and Sexual Orientation, No Qualifications or Level 1, and seven ethnicities; a smooth interaction term for IMD and VRx defined as the ratio of the Vaccine Registry divided by the Census population aged 12+; “random effects” for **LTLA** nested within **REG**. In the Bayesian formulation the parameters for “random effects” do not differ from other model parameters, all of which are estimated through their posterior distributions.

The very different scales of different covariates can make it awkward to compare their contributions. If ecdf denotes the empirical cumulative distribution function, a covariate X was first transformed to an approximately uniform distribution eeX <-ecdf(X)(X). Define nnX <-qnorm(eeX), the standard normal quantile of eeX, and put znX <-as.numeric(scale(nnX)) so znX is a monotonic transform of X to an approximately standard normal distribution with mean and sd set to 0 and 1. The various transformed covariates are still distinct. This procedure was unsuitable for a few covariates with insufficient variation.

All univariate smooth terms use the `brms` default “thin plate regression splines”[13] as in `mgcv`, with basis dimension 7 and with covariates in the zn form derived from population proportions of Census 2021 variables. The default interaction term with marginal basis dimension 5 uses the t2 tensor product of splines[25] with the zn forms of IMD and VRx. Parametric terms use the zn form, except for the military indicators and population proportions in communal defence or prisons. Most MSOA do not contain a barracks or prison and are not adjacent to a military base.

A univariate smooth term s(x,k=7) is interpreted as the sum of a linear term and 5 splines, each with associated parameters (in `mgcv` when k=7 the penalty matrix has rank 5 and a 2-dimensional subspace of parameters is unpenalized). Each of the 5 penalized spline parameters is obtained as the product of a single positive sd pertaining to the smooth term, and a standardized coefficient for the particular penalized spline. The default prior for the linear term is flat. The default prior for the sd is a half Student-t, constrained to *>* 0, with 3 degrees of freedom and scaled by 2.5. Default priors for the standardized coefficients are standard normal. An interaction term t2(x,y,k=5) is handled similarly, after decomposition into components of dimension 9, 6, and 6. Likewise, the parameters for the random effects for REG are obtained as the product of a single positive sd and standardized coefficients, with default priors as above. Parameters and default priors for the random effects for LTLA are similar. The parametric terms in the model are transformed by a QR decomposition and given default flat priors. The default prior for *ϕ* is Gamma(0.01,0.01).

Sampling used the default settings with default priors, 4 chains of length 1000 after a warmup of 1000, but maximum treedepth was reset from 10 to 13. The output was checked with standard Bayesplot tests [26] including divergence, treedepth, *R̂* (modified Gelman-Rubin statistic split-*R̂*), ebfmi (estimated bayesian fraction of missing information), posterior predictive density. Bayesian R^2^ [27] and Mean Absolute Deviation MAD = mean(abs(observed-fitted)), were computed for the fitted model.

The samples form a matrix of dimension c(4000,482). The distribution of “random effects” for a specific Region is given by the relevant column of this matrix. Forming the matrix of nine Regional columns and taking rowMeans gives the distribution of the mean Regional effect. Subtracting this distribution from each Regional column gives the distribution of each Regional effect centred by the mean Regional effect. Its mean is the estimated “centred regional effect”, and highest posterior density intervals (hdi) are obtained by `HDInterval`[28], yielding a caterpillar plot with associated hdi. To compare two regions, their columns are subtracted and a mean and hdi are obtained for the difference, which is unaffected by the centering. A similar approach was applied to plot and compare nested lower tier local authorities.

For each parametric term, mean and hdi were obtained from the relevant column. Univariate smooth terms were plotted in `brms` as conditional smooths, whose ribbons display the 0.025 and 0.975 quantiles. The interaction term t2(znIMD,znVRx) is plotted as a conditional smooth with contours added using `metR`[29]. Profiles for an individual MSOA display the 10 terms with the largest absolute contribution to the linear predictor for that MSOA. Summarising over all 6773 MSOA shows the frequency with which absolute contributions from a particular covariate are ranked 1st, 2nd etc.

### Leave-One-Out methods

A future pandemic may be very different and even if vaccines become available the pattern of uptake may also be very different. Nevertheless if new uptake data were to be generated by the same process as with COVID-19, could the fitted model predict the future? Although we do not know the true generating process for the COVID-19 data, one approach is to consider how well the model can predict the current outcome at each point from the the data at all other points. When a `Stan`model has been sampled, the programme loo (Efficient Leave-One-Out Cross-Validation and WAIC for Bayesian Models) [30] estimates the expected log predictive density (elpd) of the observed data at each point using covariates at that point and the full data at all other points. Lower values of elpd indicate failure of prediction from the rest of the data. The predicted mean value of the response at each point is also obtained from loo. The Bayesplot loo_pit_overlay tests whether it is plausible that the observed data arose from the leave-one-out distributions.

Models can be compared by the difference Δ in their respective sums of pointwise elpd. If V is the variance of the pointwise difference in elpd, estimated from the sample of pointwise differences, then sd = sd(Δ) = (*N* ∗ *V*)^1^*^/^*^2^ for N points taken as independent, and Δ / sd is tested against a standard normal distribution. The main model can be compared with models which drop a particular term or group of terms. If Δ *>* 0 is significant, the term or group is important to the model’s predictive ability. However, loo comparisons can fail. A more robust alternative `MixIS` (Mixture Importance Sampling) [31, 32], also produces pointwise estimates elpd_mixis which can be used for sd and model comparison as above. Whereas loo estimates elpd from the fitted model, `MixIS` requires a single refit using a minor modification of the code (included in Additional file 4). Terms and groups were ranked by the Δ values from `MixIS` with significance estimated from Δ / sd. When comparing models, their respective pointwise elpd_mixis values show which model fits better in any particular MSOA, and summarising this shows the frequency with which a particular model is superior. For the full model ℳ1 the predicted mean value of uptake in each MSOA was obtained from MixIS, and overall fit was measured by Mean Absolute Deviation. Possible outliers were identified using the `MixIS` estimates of pointwise elpd for the full model. With a “mixture information criterion” mixic defined as −2*elpd_mixis, high mixic values indicate points whose observed value is unlikely if the data was actually generated in accordance with the model. But how unlikely should it be before we consider the point an outlier? The programme lookout[33] identifies outliers using leave-one-out kernel density estimates and extreme value theory. It was applied to the matrix cbind(mixpred,mixic) where mixpred is the uptake predicted from the `MixIS` output, to identify exceptional points whose pointwise probability generated by lookout is *<* 0.01.

An alternative approach used the theoretical quantiles of a Generalized Pareto Distribution (GPD) fitted to the top 20% of mixic values, with scale and shape parameters estimated using fit.gpd in mev[34] with the default frequentist method “Grimshaw”. If *σ* is the estimated scale and *ξ* is the estimated shape, then for quantile U of the uniform distribution the fitted GPD has corresponding quantile *σ* ∗ (*U ^ξ^* − 1)*/ξ*. The quantiles U were chosen as the order statistics of the uniform distribution, which are beta-distributed. The U and their confidence intervals give corresponding quantiles and confidence intervals of the fitted GPD. The mixic values (above threshold) were plotted against the quantiles from the fitted GPD. Those MSOA whose mixic bor-dered or exceeded the 95% contours of the GPD fit, or was exceptionally large even though within the contours, were considered as possible outliers.

## 3 Results

Preliminary modelling identified “African & Other Black” (combined), “Caribbean”, “Pakistani”, “Roma”, “Romanian”, “Other White” and “White British” as ethnicities to include in the main model for the 1st Primary, although “Romanian” is also a category within “Other White”. Other ethnicities appeared to have less impact and were omitted to speed computation.

The main model ℳ1 for the 1st Primary passed standard tests for the functioning of the sampler. There were no divergences. The *R̂* statistic has max(*R̂*) = 1.011 and only r_REG[North.East,Intercept] has *R̂* > 1.01. Histograms of total energy *π_E_* and its stepwise differences Δ*π_E_* show good overlap, and ebfmi *>* 0.6 in all chains. The posterior predictive density plot of 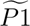/VR closely matches the observed densities.

Mean Absolute Deviation MAD = 0.011 and Bayesian R^2^ = 0.989. For the leave-one-out estimates obtained with loo or MixIS, MAD = 0.012.

Fig. 1 plots uptake against fitted uptake, where the fitted values 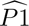 define fitted U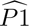 =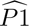/VR. MSOAs with population density < 500/km^2^ generally had higher uptake.

**Fig. 1.**
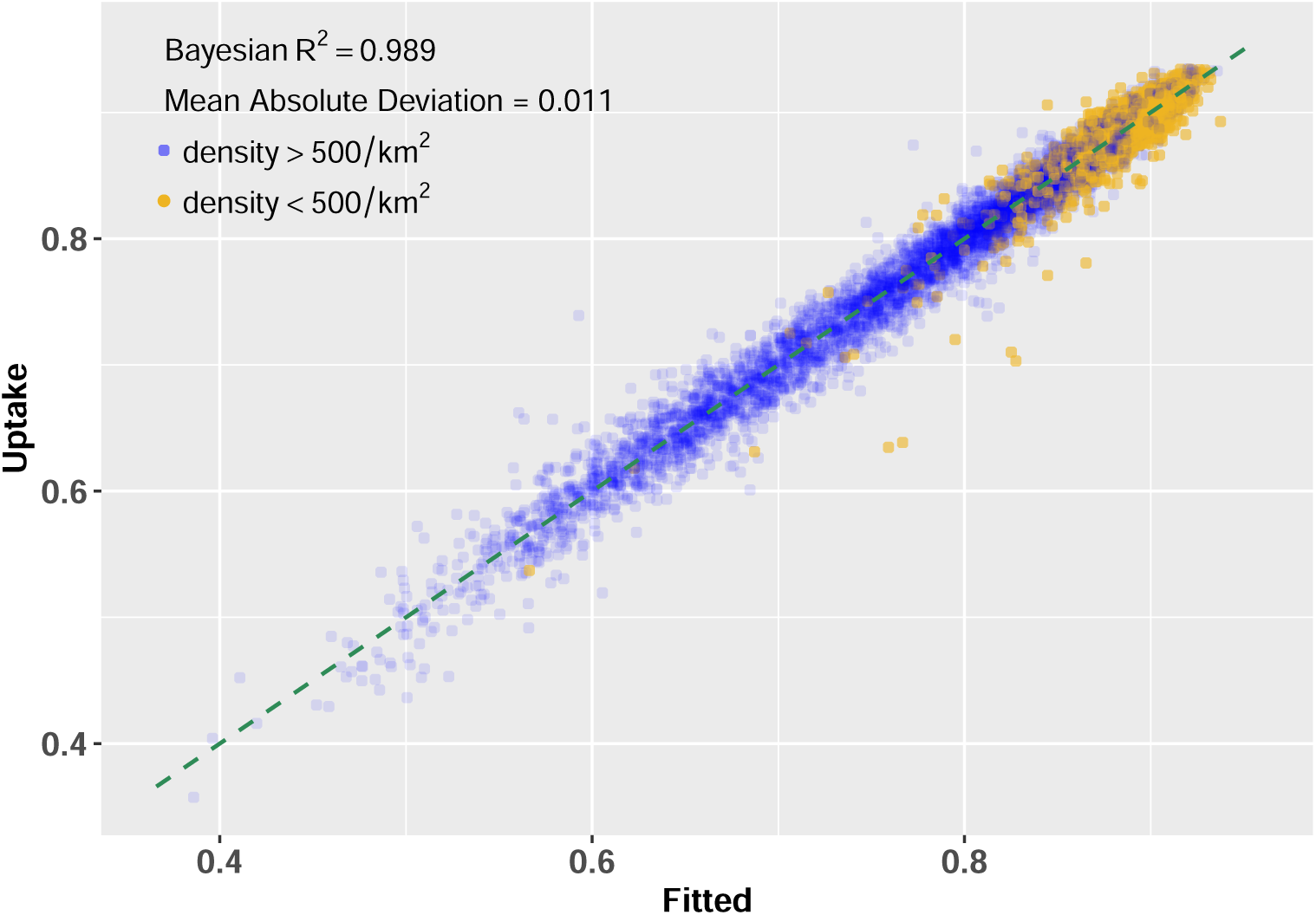
Density refers to population per km2. See Methods for Bayesian R2 and Mean Absolute Deviation.

Fig. 2 shows a caterpillar plot for the “centred regional effects” with highest posterior density intervals (hdi) of mass 0.95. London has the highest effect and North West the lowest. For London vs North West the difference has mean 0.193 with hdi (0.149, 0.244), and for London vs North East the corresponding values are 0.179 (0.121, 0.248). Whilst the centred effects for North West and North East are not credibly different, each is credibly lower than the centred effects for all other Regions. The centred effect for Yorkshire and The Humber is credibly lower than those for the East Midlands, East of England, South East, South West, and London. The centred effect for London is credibly higher than those for the East Midlands, North East, North West, West Midlands, Yorkshire and The Humber.

**Fig. 2.**
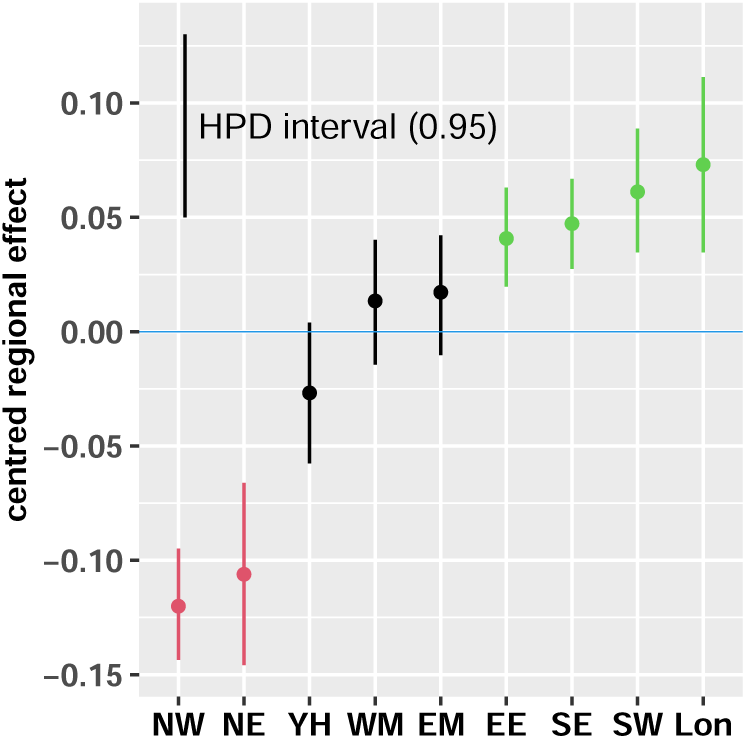
M1 (1st Primary) caterpillar plot of “centred regional effects” for the nine English regions: East Midlands (EM), East of England (EE), London (Lon), North East (NE), North West (NW), South East (SE), South West (SW), West Midlands (WM), Yorkshire and The Humber (YH). The distribution of mean regional effects is subtracted from the distribution of effects for each region. Vertical lines show the Highest Posterior Density intervals of probability mass 0.95. Colours indicate the lowest two (red) and highest four (green) centred effects.

Fig. 3 plots the random effects for LTLA nested within REG with intervals hdi, and lists the authorities with the 10 lowest and highest nested effects. The effect for Allerdale exceeds that for Solihull by 0.399 with hdi (0.316, 0.488). Differences between each of the top 10 and bottom 10 have hdi which exceed 0.

**Fig. 3.**
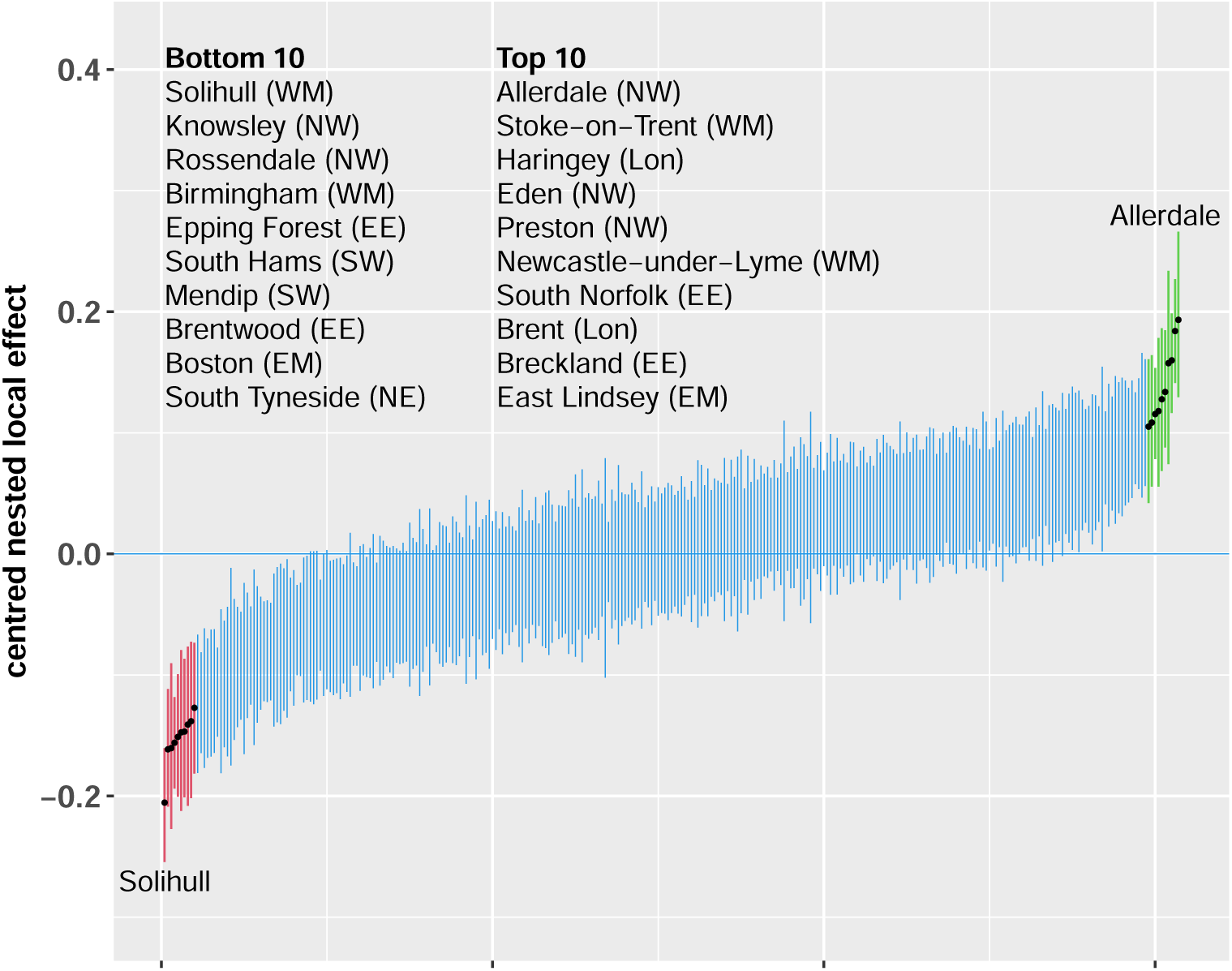
M1 (1st Primary) caterpillar plot of “centred nested local effects” for the 307 English local authorities nested within the nine regions. The distribution of mean nested local effects is subtracted from the distribution of effects for each nested local authority. Vertical lines show the Highest Posterior Density intervals of probability mass 0.95. The lowest 10 (red) and highest 10 (green) centred nested local effects are marked, and their local authorities and regions are listed.

Table 1 lists the parametric terms showing a short text description, mean and hdi. Most of the hdi exclude 0 or are borderline. The exceptions are znX711 (Sales Assistant); znX821 (Road Transport Driver); indicators for Army and Royal Navy; No Passport. The leading (absolute value) parameters are for Brexit, Finance Professional, Artist Literary & Media, Caring Personal Services, Long Term Conditions, Flu vaccination (over 65), Royal Air Force, Royal Marines. As Communal Defence Establishment and Prison are the original proportions not transformed to zn form, the corresponding parameters are much larger, with commDefence 3.881 (3.348, 4.491) and commPrison −1.317 (−1.604, −1.018).

**Table 1.**
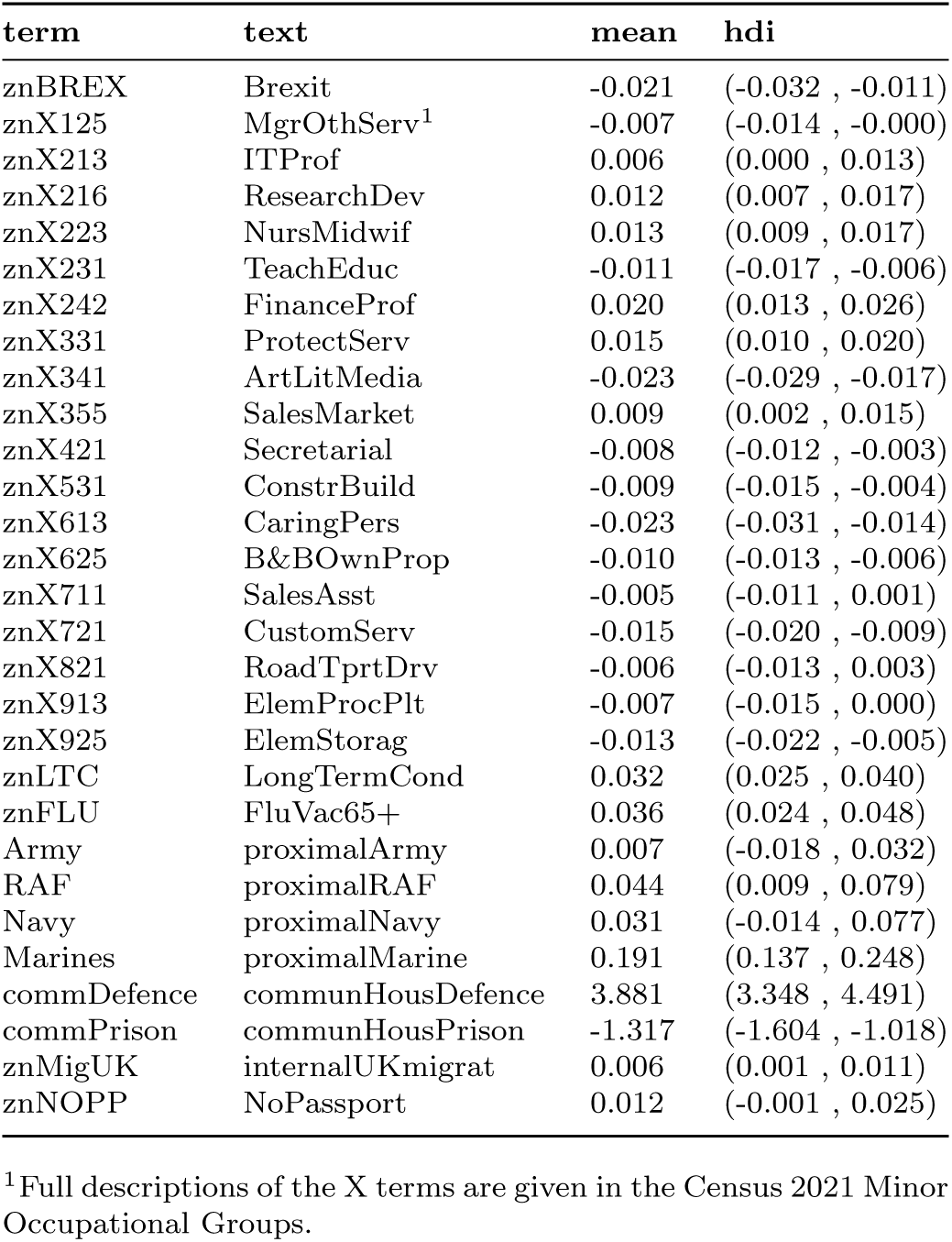
Model *M*1 parametric terms.

Fig. 4 shows the contributions to the linear predictor for the 15 univariate smoothers with ribbons for 95% credible intervals. The x-axis is labelled without “zn” to simplify the figure, and the scale for the y-axis varies. The smoothers for znAged2024 (panel **f**), znAged55+ (**g**) and znOthWh (**n**) show steeper absolute gradients than other covariates, with znOthWh the steepest.

**Fig. 4.**
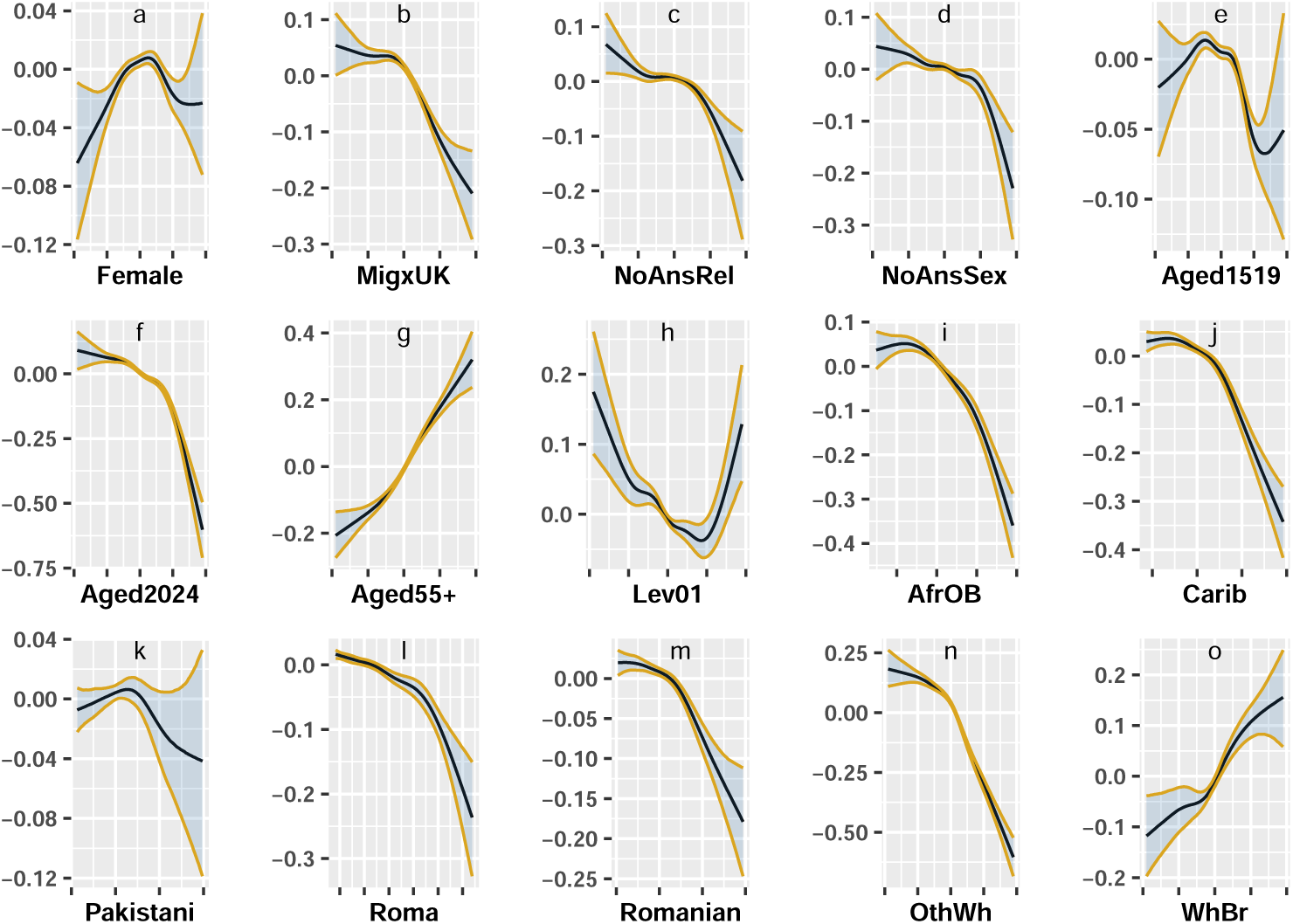
M1 (1st Primary) univariate smooth terms for scaled approximate standard normal trans-forms of covariates. Solid black curves give the mean value of the smooth at each value of the transformed covariate, whilst the ribbon boundaries show the 0.025 and 0.975 quantiles. Smooths are shown as contributions to the linear predictor rather than the linked fitted value.

Fig. 5 panel **a** shows the interaction plot for t2(znIMD,znVRx). The range of contributions to the linear predictor is greater than for any of the univariate smoothers. Harehills South has an interaction contributing −0.383, while Ormskirk Town & East contributes 0.632. Panel **b** shows the fitted effects of znIMD when conditioning on znVRx, and panel **c** shows the fitted effects of znVRx when conditioning on znIMD.

**Fig. 5.**
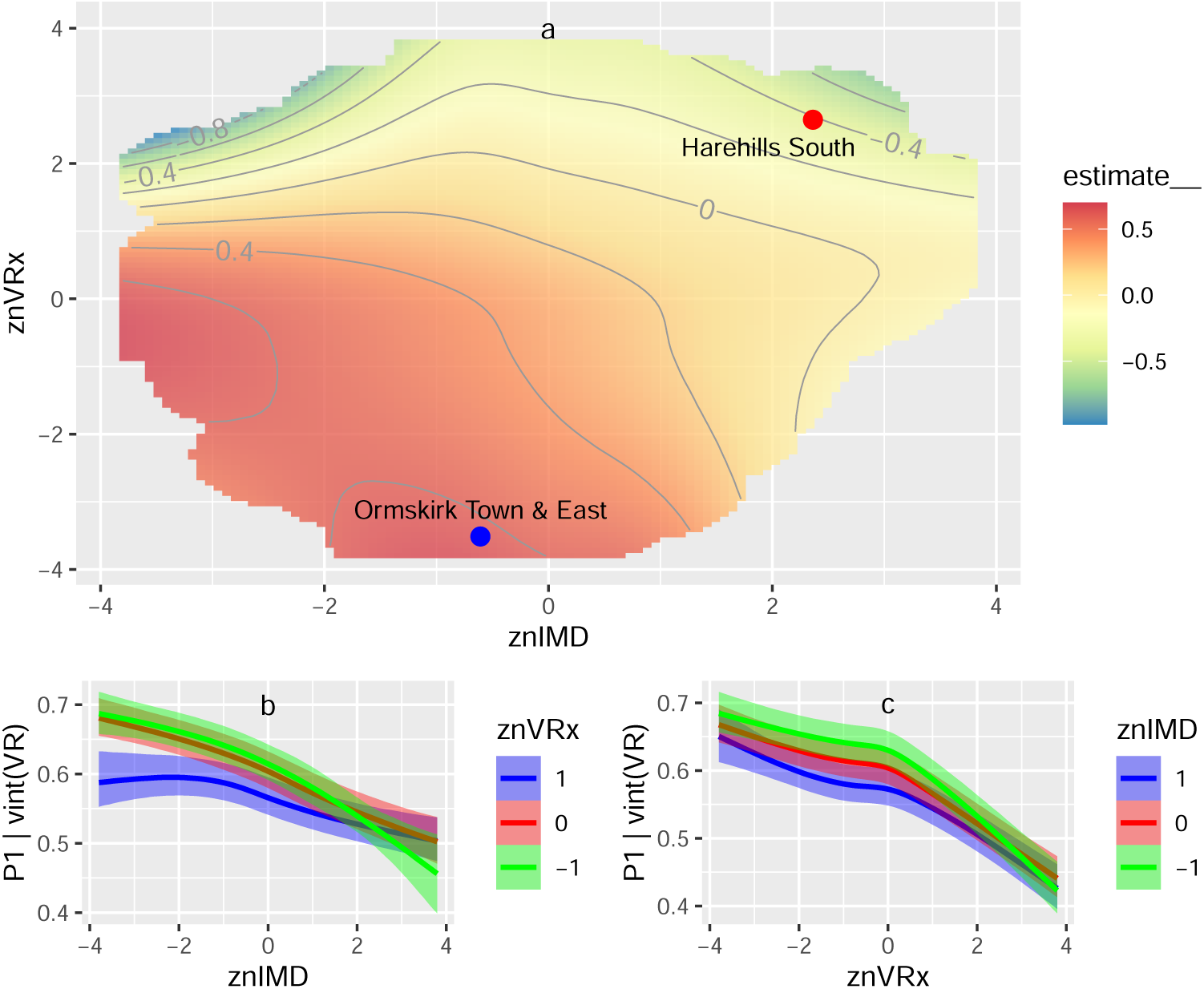
Panel a: Bivariate smooth for the interaction of transformed Index of Multiple Deprivation and the Vaccine Registry divided by the Census population aged 12 and over. The prefix zn denotes the transform to an approximate standard normal distribution. Contours show the contribution to the linear predictor, not the linked fitted value. The blue and red points illustrate the difference between high (blue) and low (red) contributions. Panel b: Fitted value attributed to varying znIMD if znVRx is fixed at 1, 0, or −1 and all other terms contribute 0 to the linear predictor. Panel c: As with Panel b but varying znVRx with znIMD fixed.

Codes for generating Figs. 1-5 are given in Additional file 5.

Fig. 6 shows the profile plot for Harehills South, the MSOA with the lowest uptake in England. Panel **a** identifies the MSOA within its local authority and region, and reports its uptake for the 1st Primary. The plot shows the leave-one-out predicted values from `MixIS` against uptake for all MSOA, and highlights those in the local authority (yellow) and the MSOA itself (red). Panel **b** identifies the 10 covariates with the largest absolute contribution to the linear predictor, in descending order and showing their hdi. Whilst znLTC (Long Term Conditions) is a positive predictor it makes a negative contribution here as znLTC = −2.013 for Harehills South. Contributions for LTLA and REG are centred by their means (REG is not among the top 10 for this MSOA) and labelled as LTLAc and REGc if they occur. Panel **c** places the MSOA on the interaction plot t2(znIMD,znVRx). The remaining panels place the MSOA on the smooth curves, or straight lines for parametric terms, for the top five such terms identified in panel **b**, with ribbons showing 95% credible intervals. Indicator variables for proximity to military bases are not plotted though they may appear in panel **b**. Code for generating Fig. 6 is given in Additional file 6.

**Fig. 6.**
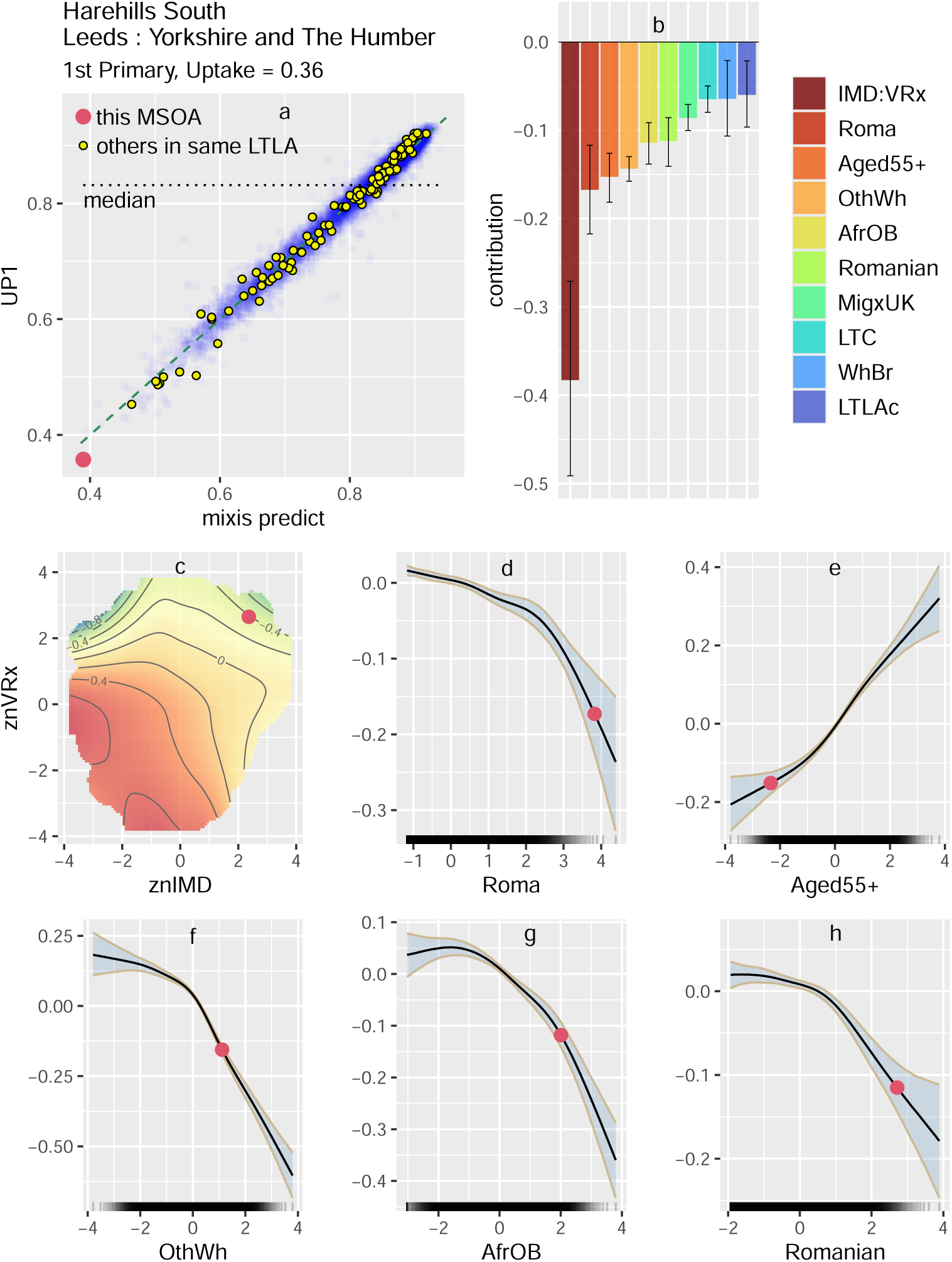
MSOA profile plot for Harehills South 1st Primary. See main text (Results) for details

Fig. 7 Panel **a** shows the plot of mixic vs the `MixIS` predicted values, with possible outliers identified by lookout. In descending order starting with the most exceptional (lowest probability), these are Fratton West & Portsea, Soham, Abbey Field, Boughton & Selling, Leake & Butterwick, Forest Row & Coleman’s Hatch, Salford Central & University. Panel **b** plots the observed and theoretical quantiles of a Generalized Pareto Distribution for the top 20% of mixic values, and identifies the same points as exceptional. Panel **c** plots uptake against the `MixIS` predicted values, and marks the possible outliers. Code for generating Fig. 7 is given in Additional file 7.

**Fig. 7.**
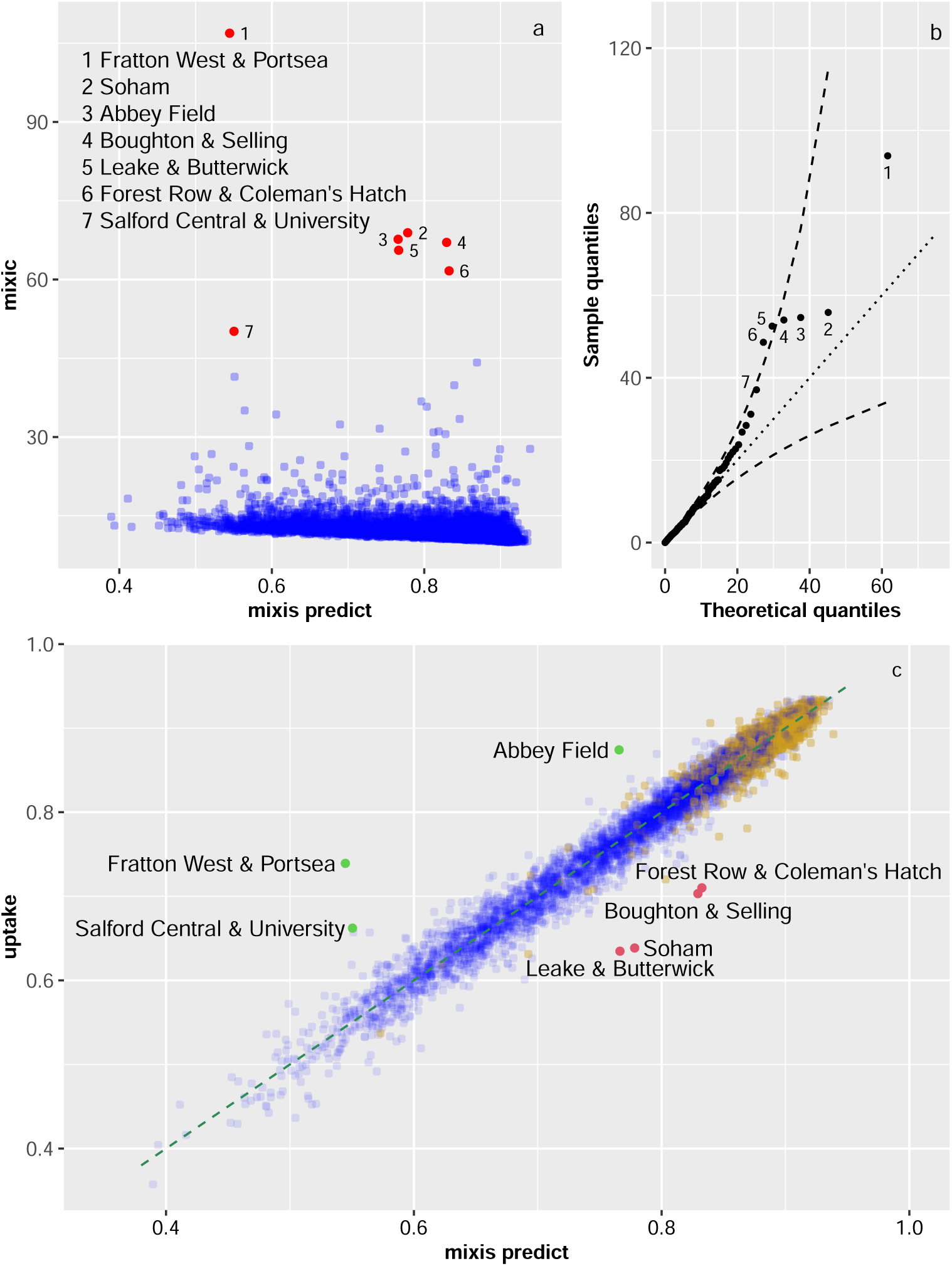
Possible outliers for 1st Primary. See main text (Results and Methods) for details

Table 2 shows the impact of deleting selected terms and groups from the model. For each term Δ, its sd and the resulting two-sided p value are obtained as in Methods, par shows the number of parameters in the term, and dpar shows Δ/par, the change in summed mixic per parameter. The bold rows refer to terms which do not appear as such in the main model. The row with term (1|LTLA) refers to the comparison in which (1|REG/LTLA) is replaced by (1|REG). The row with term (1|REG) is analogous. The row with term s(znVRx) refers to the comparison in which t2(znIMD,znVRx) is replaced by s(znIMD,k=7). The row with term s(znIMD) is analogous.

**Table 2.**
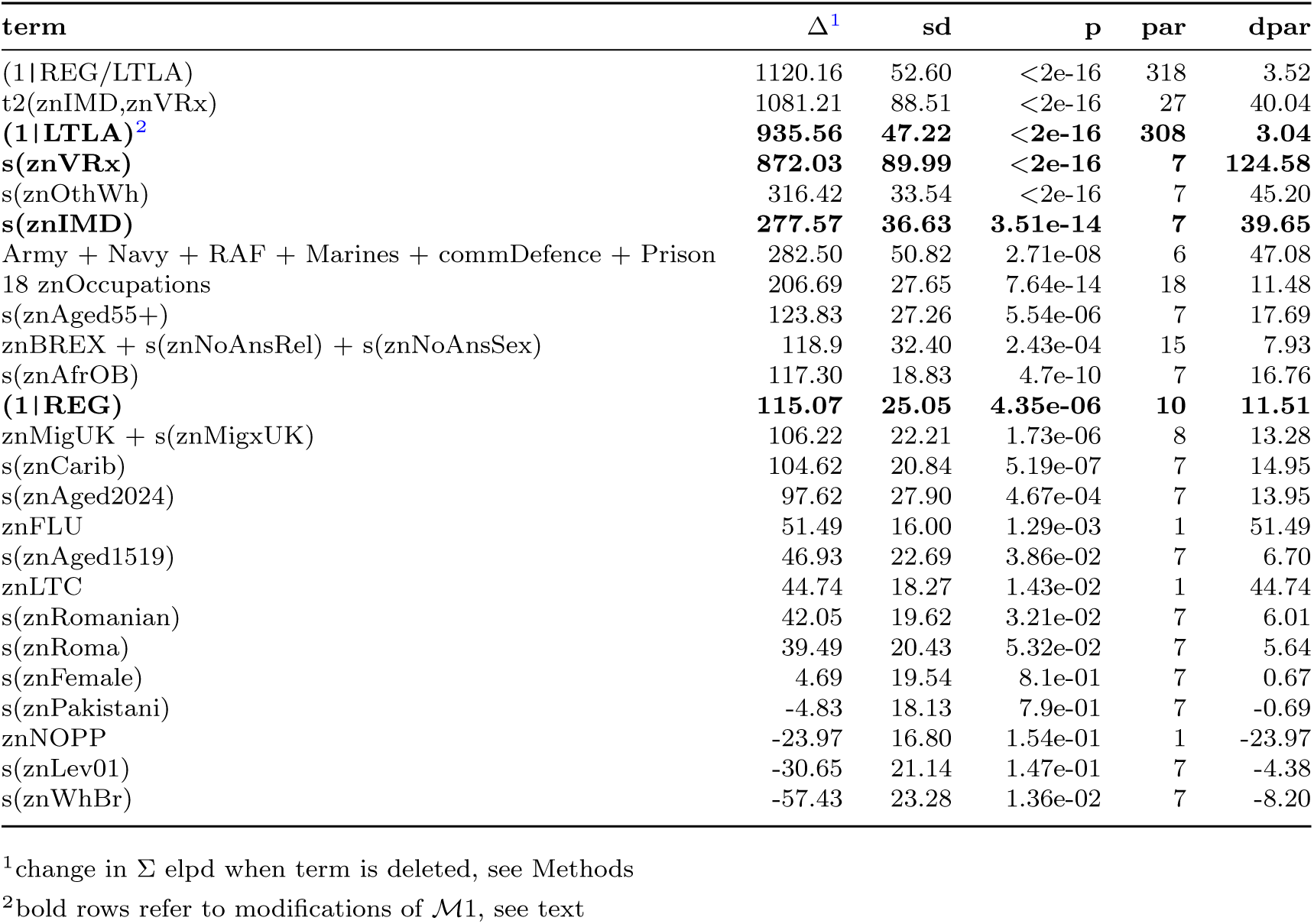
Model *M*1 Impact of deleting selected terms.

The nested factor term (1|REG/LTLA) has the highest Δ, but dpar is low as the term involves 318 parameters (9 regions + 307 local authorities + 2 sd for the random effects). The term (1|LTLA) has much larger Δ than (1|REG), but involves many more parameters. For 76% of MSOA the main model fits better than the model in which (1|REG/LTLA) is dropped, and for 74% if it is replaced by (1|REG), *i.e.* dropping the LTLA component, but only 57% if it is replaced by (1|LTLA).

The interaction term has the second highest Δ, but with only 27 parameters dpar is much higher, and znVRx is the dominant component. The interaction term made the largest absolute contribution to the linear predictor in 77% of MSOA.

Of the terms in the model (rather than separate components of the nested factor or the interaction), the univariate smoother for znOthWh has the third highest Δ. It made the second largest absolute contribution to the linear predictor in 32% of MSOA and the third largest in 23%.

Fig. 8 panel **a** plots the ℳ1 elpd estimates from loo against those from MixIS, showing small differences. Code for generating Fig. 8 is given in Additional file 8. Panel **b** plots the ℳ1 predicted mean values from `loo` against those from `MixIS`, showing very little difference. Panel **c** is the loo_pit_overlay plot for ℳ1, showing some departure from uniformity. Panel **d** is the corresponding plot for M3, defined below.

**Fig. 8.**
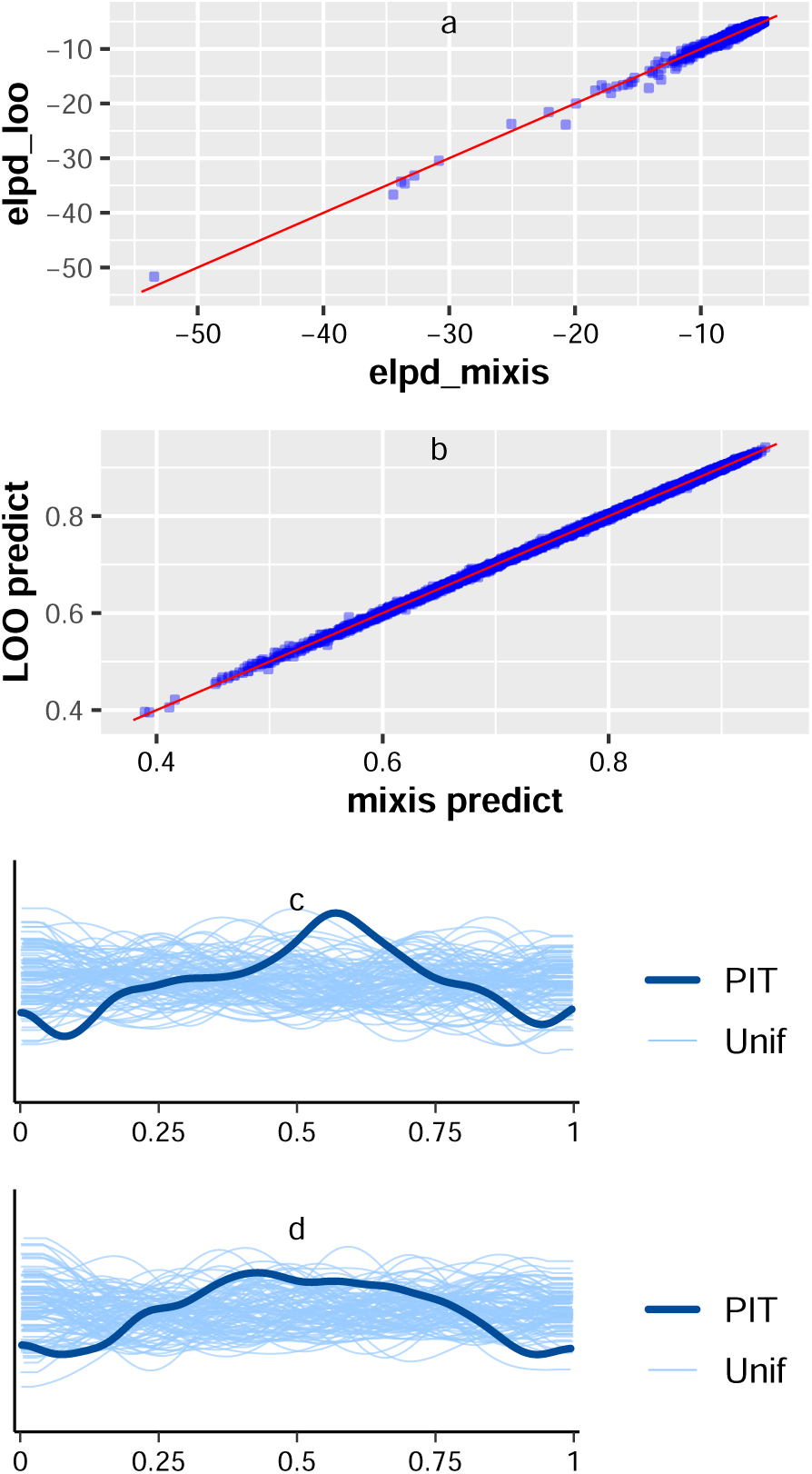
loo vs MixIS, and loo-pit-overlay plots. Panel a plots the M1 elpd estimates from loo against those from MixIS. Panel b plots the M1 predicted mean values from loo against those from MixIS. Panel c is the loo-pit-overlay plot for M1. Panel d is the corresponding plot for M3.

The uptake of the response pair (P2,VR-P2) can be modelled effectively using a single smoother s(UP1,k=7) to define the beta_binomial model ℳ2 with no other covariates. It has MAD = 0.005 and Bayesian R^2^ = 0.998. In turn, 2nd Primary uptake UP2 was a key predictor for the uptake of B3I. Model ℳ3 updated ℳ1 with the addition of s(UP2,k=7), and used the response pair (B3I,VR-B3I). The specification of ℳ3 and code for fitting it is included in Additional file 9. Fitting the model with 4000 samples had one divergence, passed standard Bayesian checks including loo_pit_overlay (see Fig. 8 panel **d**), MAD = 0.006 and Bayesian R^2^ = 0.997. The UP2 term made the largest absolute contribution to the linear predictor in 93% of MSOA.

Fig. 9 shows the univariate smoothers for M3. Code for generating Fig. 9 is given in Additional file 10. The curve for UP2 (panel **a**) rises steeply from below −1.5 to over 0.5. The curve for znAged55+ (**h**) is similar to its appearance when modelling P1 (Fig. 4 **g**). However, the curve for znAged2024 (**g**) differs from its appearance when modelling P1 (Fig. 4 **f**). Likewise the curve for znOthWh (**o**) differs markedly from (Fig. 4 **n**), and the curve for znPakistani (**l**) differs markedly from Fig. 4 **k**.

**Fig. 9.**
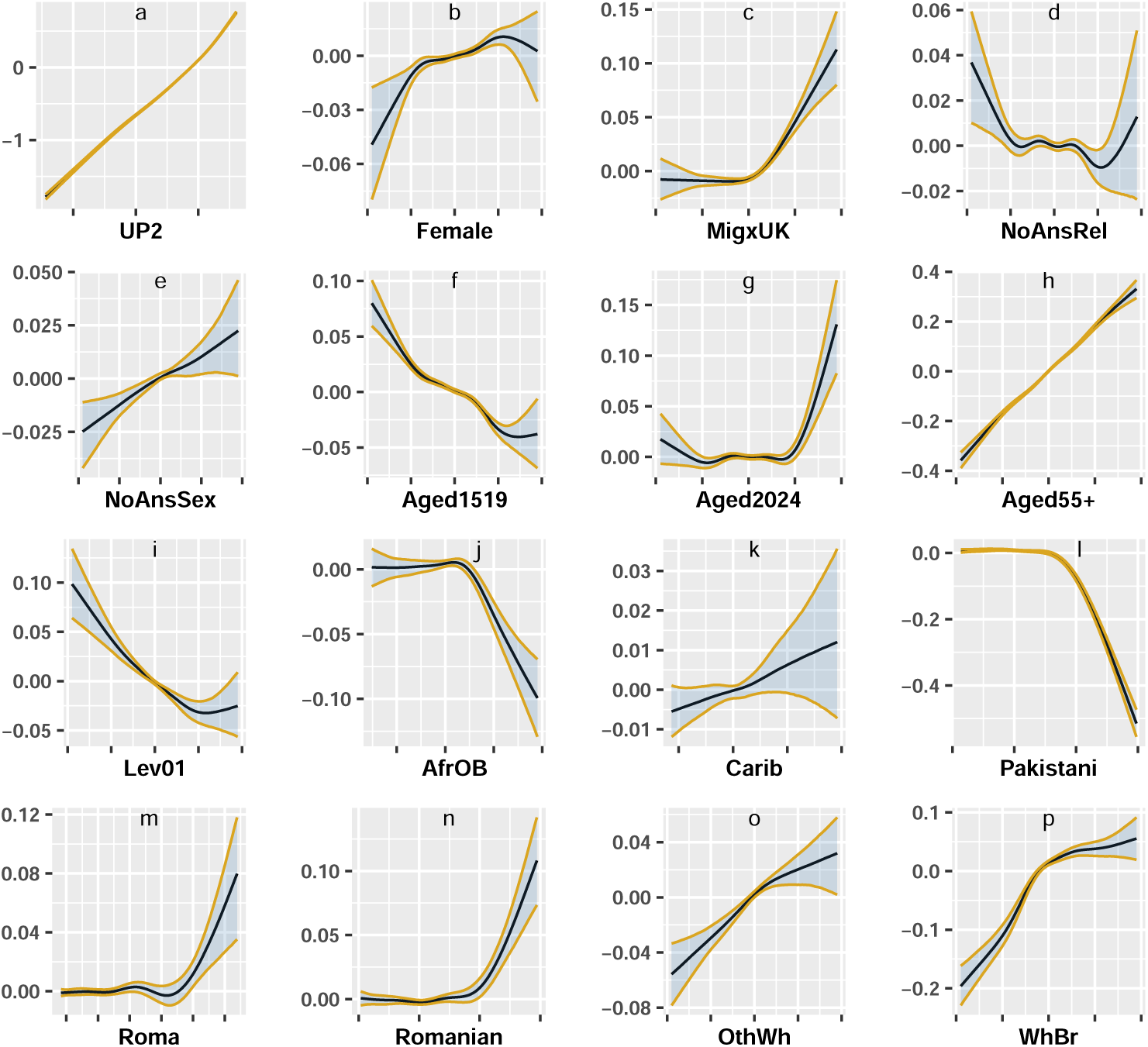
M3 (Booster / 3rd Injection) univariate smooth terms for uptake of 2nd Primary (UP2) and scaled approximate standard normal transforms of other covariates. Solid black curves give the mean value of the smooth at each value of UP2 or transformed covariate, whilst the ribbon boundaries show the 0.025 and 0.975 quantiles. Smooths are shown as contributions to the linear predictor rather than the linked fitted value.

The parametric intervals, a plot of uptake vs fitted values and caterpillar plots for REG and LTLA (nested) using ℳ3 along with the codes for generating these figures are Additional files 11-15. znBREX has negative impact −0.008 (−0.012, - 0.004). London has the highest centred regional effect and North West the lowest, with difference 0.041 (0.022, 0.060). The centred regional effect for North West is credibly lower than those for all other regions except North East. The London effect is credibly higher than those for the East Midlands, North East, North West, and West Midlands. For the Local Authority nested effects, Rushmoor (highest) vs Blackburn with Darwen (lowest) = 0.171 (0.141, 0.201). Differences between each of the top 10 and bottom 10 have hdi which exclude 0. The only possible outlier identified by both `lookout` and mev is Batley Carr & Mount Pleasant in Kirklees, though `lookout` also finds less extreme possible outliers Belgrave South, Nelson West, and Crowthorne South.

Fig. 10 shows the B3I profile for Soham, a possible outlier when modelling P1. Code for generating Fig. 10 is in Additional file 16. For B3I the uptake 0.5 is just above the fitted value 0.494 and the `MixIS` prediction 0.493. UP2 has the largest impact on the linear predictor for Soham (panel **b**). The parametric term for No Passport (**b f**), which was a positive but not quite credible predictor for P1, has a stronger additional positive credible impact on B3I.

**Fig. 10.**
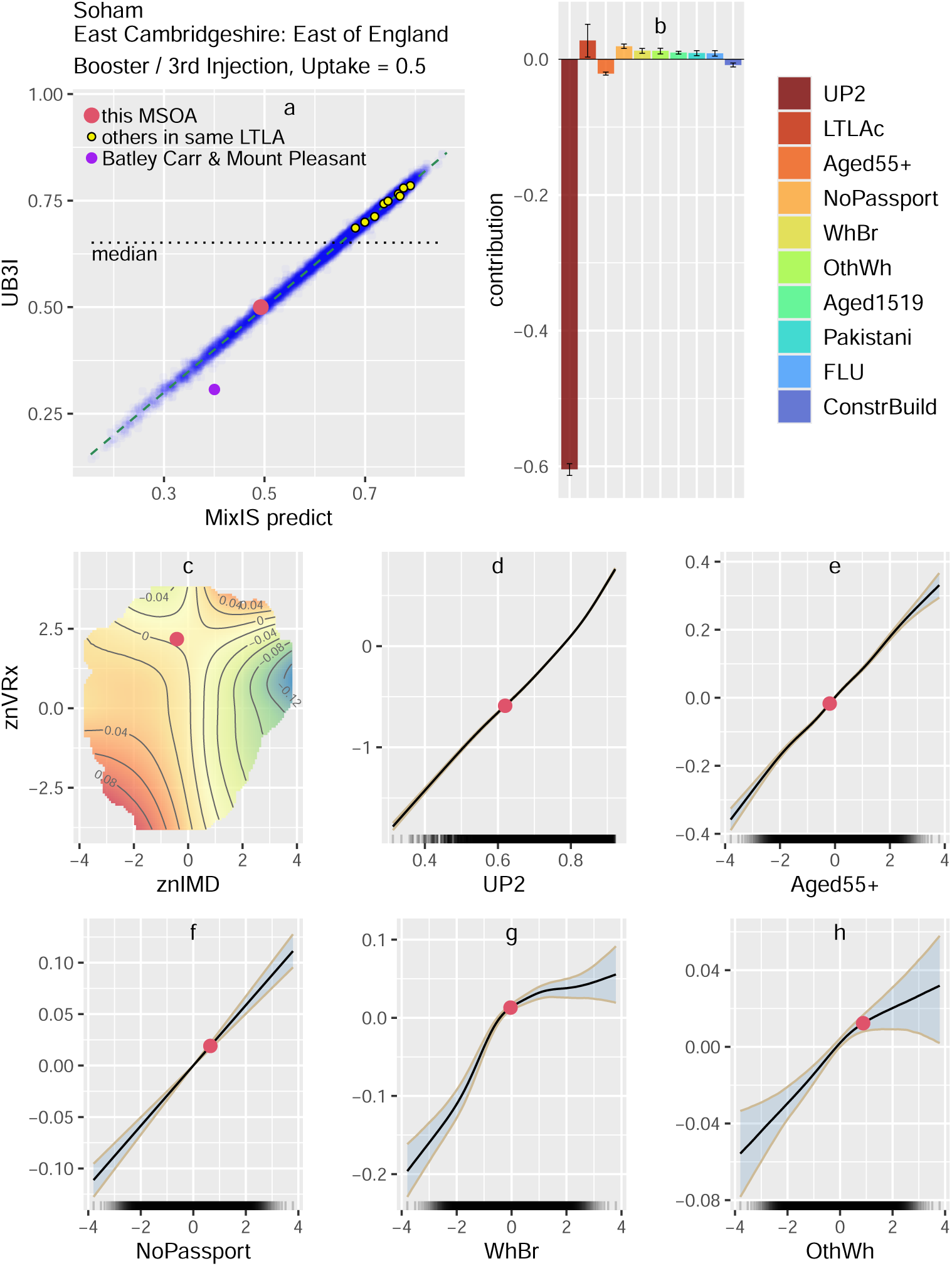
MSOA profile plot for Soham Booster / Third Injection. See main text (Results) for details

## 4 Discussion

Variation in uptake has real clinical consequences in view of strong evidence that the vaccines were effective in England in reducing mortality and the need for critical care in hospital, although effectiveness waned over time[35–37]. The results here show that uptake of the 2nd Primary was almost completely predicted by the 1st Primary, and in turn the 2nd Primary uptake was the most important predictor for the Booster / 3rd Injection. Therefore, the analysis focused on the 1st Primary. Whilst individual records are essential for clinicians, grouped area data can help in understanding why vaccine uptake varies.

Bayesian models succeed in explaining the uptake at Middle Super Output Area level. Fitted values and leave-one-out predictions from `MixIS` or loo are generally close to the observed values. The models pass tests for convergence, Mean Absolute Deviations are low, and values of R^2^ are close to 1. Only seven MSOA (0.1%) were identified as “possible outliers”, whose uptake was exceptional if the model corresponded to the actual process by which data was generated.

As the models fit the overall data, the contribution of terms to the fitted values are estimates of how their covariates impact on the uptake itself. Profile plots show the leading contributions to the linear predictors for a particular MSOA, and the response curves for the five largest (positive or negative) contributors. A covariate with a steep response curve can discriminate between different MSOA, but its impact for a particular locality depends on the MSOA’s position on the curve. Profiles vary widely for different MSOA.

If people entered or left the MSOA but the Vaccine Registry (VR) was not adjusted accordingly, the denominator could misrepresent the eligible population causing mis-estimates of the true uptake. NHS England recognised at the outset that National Immunisation Management System (NIMS) denominators used for VR were not always accurate but still recommended using them to estimate uptake.[38] I set VRx as the ratio of VR to Census population aged 12+. If the Census was accurate and the population stable with high levels of registration, we might expect VRx ∼ 1. In fact VRx ranges from 0.62 in Lakenheath to 2.30 in Fitzrovia West & Bloomsbury East. With mean 1.12 VRx is rarely below 1 and around 5% of values exceed 1.35. Both VRx and IMD are highly credible predictors of uptake, with a strong interaction. Modelling with only the interaction term was faster and nearly as powerful, so univariate smoothers for VRx and IMD were omitted. The interaction made the largest absolute contribution to the linear predictor in 77% of MSOA. In model comparisons (Table 2) the impact of dropping the interaction was almost as large as dropping the factor term for local authority nested within region, with a much higher impact per parameter.

In December 2020, SAGE[2] anticipated “Black ethnic groups [are] the most likely to be COVID-19 vaccine hesitant (28% reporting intention to vaccinate), followed by the Pakistani/Bangladeshi group. Other White ethnic groups (which include Eastern European communities) also had higher levels of COVID-19 vaccine hesitancy than White UK/White Irish ethnicity (high confidence)”. As of July 2021 the monthly data published by NHS England[39] showed that for people aged 50+ the uptake of two doses was lowest for Black Caribbean (59.5%), followed by Other Black (60.8%) and Black African (63.1%) whilst uptake for the Other White group was 76.1%. Later individual modelling by Gaughan et al.[6, Fig. 2] which adjusted for demographic, socio-economic and health covariates found that the Other White group was second to Black Caribbean for the likelihood of not receiving a vaccine by 15 June 2021. Whilst SAGE anticipated high levels of hesitancy in the Bangladeshi community, this was not shown in Gaughan et al. This may reflect the intensive efforts of the community itself to promote the vaccine.[40, 41]

To simplify visual comparisons and aid convergence most covariates were monotonically transformed to approximately standard normal distributions, and this included all the covariates in univariate smooth terms and the tensor product interaction. From Fig. 4, the transformed “Other White” covariate (znOthWh) was a negative predictor with a steeper absolute gradient than any other univariate smooth term. This ethnic category conveys more information about the predicted uptake in a particular MSOA than the age groups 20-24 or 55+, but all three are more informative in this sense than other univariate terms.

When considering the impact of dropping individual terms or groups on overall predictive ability (Table 2), the “Other White” term ranked third, behind the interaction term and the nested factors for region and local authority, but far above other ethnicities, though most of these were still significant. Before rollout of the 1st Primary, de Figueiredo [9] had identified Polish speakers as a group with negative intention to vaccinate, later validated with data on uptake as of 2021.

Area impact of an ethnicity depends on its distribution within the MSOA as well as individual response. The relatively small subgroup “Black Caribbean” with very poor individual uptake had less impact on MSOA uptake in general than the much larger subgroup “Other White”. The largest subgroup “White British” was a positive predictor (Fig. 4 panel **o**) but its overall impact was negative (Table 2). The model excluding “White British” was better able to predict the overall uptake, suggesting that during model fitting other terms in the simpler model adjust to compensate for the missing covariate.

As expected, aged 55+ is a strong positive predictor for P1 (Fig. 4 panel **g**), ranking sixth in model impact, and was an additional positive predictor for B3I when controlling for UP2 (Fig. 9 panel **h**). The proportion aged 20-24 is a strong negative predictor for P1 (Fig. 4 panel **f**).

SAGE anticipated that distrust would affect uptake, and this was confirmed in a systematic review of 24 UK studies on vaccine hesitancy among ethnic minorities[42] and a study of barriers to COVID-19 vaccine uptake in London.[43] Distrust in government is not limited to ethnic minorities. Three model terms may indicate it: declining to answer the Census questions on Religion and, separately, on Sexual Orientation, and the “Leave” vote in the 2016 EU Referendum.

Declining to answer each of the two Census questions was a credible negative predictor (Fig. 4 panels **c**, **d**). The “Leave” data is only available at local authority level and was projected to apply to each MSOA within the LTLA, an unlikely oversimplification. Nevertheless, the parametric term znBREX was a credible negative predictor (Table 1). In model comparisons (Table 2), the “distrust group” ranked seventh. When modelling B3I controlling for UP2, znBREX was again a negative predictor. Thus the Brexit vote was associated with lower uptake of P1 with concomitant impact on UP2, and then had a further negative impact on uptake of B3I. Having no passport was correlated with Brexit, but had a positive impact on P1 and additionally on B3I. The “Brexit” effect was analysed by Phalippou and Wu[44] who found the percentage voting to Leave the EU or abstaining was a strong negative predictor of uptake, particularly for the Booster.

Internal UK Migration is a positive predictor for P1 (Table 1) whilst International Migration is strongly negative (Fig. 4, panel **b**). The group comprising internal and international migration ranked ninth. Evidence to the UK COVID-19 Inquiry[1, 7, 8] highlighted specific barriers to vaccine uptake for international migrants.“Profoundly low” GP registration was linked to systematic, and illegal, refusals to register patients with insecure immigration status or lacking proof of right to residency. Patients did not always know that COVID-19 vaccination was freely available. The Hostile Environment, with charges for secondary care and data-sharing between the NHS and the Home Office with a risk to visa applications and possible deportation, led some migrants to fear any contact with the NHS. Although the UK Government announced in February 2021 that GP Registration and access to the vaccine did not depend on immigration status, there was no statement ruling out data-sharing or immigration enforcement as a result, and the fear remained.

Eighteen minor occupational groups were selected from the preliminary modelling with `mgcv`, and most were credible predictors in ℳ1. Of these, the largest positive predictors were “Finance Professionals” and “Protective Service Occupations” (including Non-Commissioned Officers, Police Officers, Fire Service Officers, Customs and Border Officers), whilst the largest negative predictors were “Artistic, Literary and Media Occupations” and “Caring Personal Services” (including nursing auxilliaries, ambu-lance staff excluding paramedics, dental nurses, care workers and home carers). The combined group ranked fifth for overall impact.

Military bases and prisons had a strong impact on uptake of P1 (Table 1). As family housing can be sited in an MSOA adjacent to the base, indicators were defined for MSOA which contain or are adjacent to a military site. For P1 uptake, proximity to a Royal Marines base was a large, credible positive predictor, although this involved only 19 MSOA, and proximity to a Royal Air Force base (64 MSOA) was also a credible positive predictor. The proportion housed in a communal defence establishment (139 MSOA) was a strong, very large positive predictor, whilst prison (97 MSOA) was a strong negative predictor. Despite the relatively small number of MSOAs affected, the impact of the combined group ranked fourth.

### Regional and Local Random Effects

The raw data shows regional differences with lower uptake in London. However, the “centred regional effect” of London in the context of other covariates is higher than those of any other region, and credibly higher than those of the East Midlands, North East, North West (lowest), West Midlands, Yorkshire and The Humber. The uncentred London “regional effect” represents the difference between the predictions for MSOA in London, and what would be expected from all other terms apart from Region. If an MSOA and its Lower Tier Local Authority were relocated from the North West to London leaving all other model covariates unchanged, the linear predictor would rise by 0.193, and a fitted uptake of 0.5 would give a new fitted uptake 0.55. This contrasts with reports which present unmodelled regional data. An ONS report in August 2021 stated that “The rate of not receiving a vaccine among over-50s is twice as high in London as anywhere else” without commenting on whether this was to be expected in view of London demography and deprivation.[45] Evidence to the COVID-19 Inquiry regarding Vaccine Hesitancy from the London School of Hygiene and Tropical Medicine (LSHTM)[1, p64] describes London as having “higher hesitancy than other parts of the UK” without comment on London demography, but refers elsewhere to modelling by de Figueiredo [9] which suggests reasons why London uptake was lower than expected from pre-rollout surveys of intention.

Local authority effects had a large overall impact. For example the London borough of Haringey had uptake 0.64, far below the national median 0.83, but its “centred nested local effect” was 0.16 (0.12, 0.20), the third highest in England. Haringey has a younger, ethnically diverse population and high levels of VRx suggesting rapid turnover. The uncentred Haringey “nested local effect” is the difference between the fitted uptake for MSOA in Haringey and what could be expected for them from all the other model covariates, including Region. For the local effects, the effect of centering is minimal.

The ten highest and ten lowest local effects are highlighted in Fig. 3. When comparing each of the highest with each of the lowest, the differences are credible as the highest posterior density intervals (95%) are strictly positive. These credible differences may reflect unmodelled covariates which could be other demographic or economic variables, or could involve the functioning of the NHS and public health departments. Allerdale and Solihull, with the highest and lowest local effects, suggest one possibility. Allerdale in North Cumbria had twelve MSOA with mean uptake 0.893, and seven had uptake *>* 0.90. The population (96,200) is overwhelmingly older, with 40% aged 55+ (against 31.6% for the North West). Ethnicity is 96.7% White British (against 81.2% for the North West), and all other ethnicities included in the model are marginal except Other White (1.4%, against 3.4% for the North West). The Vaccine Registry ratio VRx is near 1. Each of these model terms raises the fitted values. Separately, the large positive Allerdale “random effect” may reflect unmodelled covariates. Car ownership was generally high, with only 18.5% of households having no car or van (24.7% in North West), but 34.9% in Workington West. In March 2021 vaccine distribution comprised four GP surgeries, with a pharmacy added in April 2021. By October 2021 there were three GP surgeries and four pharmacies, and four of these sites were in Workington.

The NHS made vigorous efforts to promote vaccination in Allerdale. The Director of Public Health Colin Cox pointed to “Primary Care Networks in the Allerdale area both being exceptionally proactive and having a very receptive population”. (personal communication) The North Cumbria Primary Care Alliance Annual Report 2021/2022 mentions PCN involvement in vaccination clinics and sites in Workington, Carlisle and Copeland.[46] Susan Mann of the System Vaccination Operations Centre said the North Cumbria PCNs “support the vaccination campaigns with a range of clinics and both care home and housebound visits. North Cumbria is also a high performing area for uptake of all the mainstream vaccination campaigns so population acceptance of vaccines will be a contributing factor.” (personal communication) The Primary Care Services North Cumbria Team mentioned “well-established prac-tice–patient communication channels, engagement with vulnerable patient groups, and strong public messaging, rather than any unique strategies implemented by the PCNs themselves”. (personal communication) The “random effect” for the adjacent local authority Eden, also a large mainly rural area, is ranked 4th highest.

Solihull in the West Midlands had 29 MSOA with mean uptake 0.82, and six had uptake *<* 0.75. The population (216,245) is slightly older, with 34.2% aged 55+ (against 31.1% for the West Midlands) and 77.9% are White British. The low uptake areas had relative large Black Caribbean communities. In March and April 2021 vaccine distribution comprised four GP surgeries and one hospital hub. By October 2021 two pharmacies had been added. The number of distribution sites was comparable to Allerdale, but Solihull has over twice the population. Car ownership was high, with only 17.6% of households having no car or van (21.5% in West Midlands), but each of the six MSOA with low uptake had over 25% with no car or van. Primary Care Networks were active in the vaccination programme.

The contrast in “random effects” for Allerdale and Solihull is not an artefact of the interaction term or even the inclusion of znVRx in the model. Replacing the interaction by a univariate term for znIMD weakens the fit considerably, but with that simpler model eight of the previous highest (lowest) ten “random effects” are still in the highest (lowest) ten. The “random effect” for Eden using the simpler model is highest, Allerdale is second highest, whilst Solihull remains lowest.

National financial support does not explain the difference. In Allerdale, the COVID-19 Additional Funding, Contain Outbreak Management Fund and other national financial support during 2020-21 and 2021-22 totalled £4.77m. In Solihull, the corresponding figure totalled £59.73m.[47]

Across England, including in Solihull, NHS and Public Health staff made great efforts to distribute the vaccine. However, one factor which may contribute to the “random effect” is the level of communication with communities whose main language is not English. In Allerdale and Eden the only non-English main languages spoken by over 100 people are Polish (Allerdale 211, Eden 324) and Romanian (Allerdale 124, Eden 395). The Solihull population includes 9,424 people whose main language is not English, including French (260), Portuguese (245), Spanish (243), Polish (719), Romanian (492), Arabic (453), Farsi (178), Urdu (664), Hindi (262), Panjabi (1,163), Bengali (245), Gujarati (665), Telugu (317), Tamil (411), Cantonese Chinese (364), Other Chinese (431). The 1,156 Solihull residents who were migrants from outside the UK in the year before the Census may have faced language barriers as well. Yet in 2021 neither the Annual Report of the Solihull Director of Public Health nor the Solihull Health and Wellbeing Board agenda packs[48, 49] mention language barriers to vaccination, or provision of translation or interpreters. Community Champions were recruited but their Induction Briefing does not mention language.[50] An evaluation of the Community Champions programme in three West Midlands local authorities (including Birmingham but not adjacent Solihull) noted that the Vaccine Toolkit was not translated.[51]. In Birmingham, with the fourth lowest “random effect”, the Health and Wellbeing Board papers for January 2021 mention “making sure translated materials were available in people’s first languages” but the COVID-19 Inequalities Overview in March 2021 (wrongly titled March 2020) does not mention language when discussing vaccination.[52, 53]

By contrast, in Haringey the March 2021 Health and Wellbeing Board had an extended discussion of overcoming language barriers to vaccination, including GPs communicating in multiple languages.[54, p9] In Brent, with the eighth highest “random effect”, the April 2021 Health and Wellbeing Board highlighted community translation videos filmed by NHS workers in 11 community languages. In July the Brent Board mentioned different language versions of the Vaccine Bus asset, including in Romanian and Portuguese.[55] The Solihull and Birmingham reports for 2021 do not mention a Vaccine bus.

I was unable to find data on the provision of interpreters during the 1st Primary. A rapid systematic review of uptake in UK minority ethnic groups by Kamal et al.[4] stated “Lack of access to information also resulted in communication barriers largely due to low health literacy, poor other language provision, and increased digitalisation of communications. This was particularly an issue for migrant groups due to lack of access to, or knowledge of, technology.” LSHTM [1] refer to the failure of central government messaging on vaccine safety to reach various communities, as it was only delivered in English, and “For migrants attempting to seek care and information during the pandemic, language and communication issues were persistent barriers and often served to compound fear and hesitancy.” A qualitative study by Knights et al.[56] interviewed healthcare professionals and migrants in 2020 before rollout. One GP told researchers “I think the biggest problem [for COVID-19 vaccine uptake] is going to be language and culture… The big issue, basically, is language getting out there to people who need it. The people that need it, it probably won’t get to them because they’re not interacting with their health necessarily in the way that the healthcare system was built to do…”

### Outliers

Soham in East Cambridgeshire is one of seven possible outliers for the 1st Primary (Fig. 7), with uptake 0.64 and leave-one-out (MixIS) prediction 0.78. The Census shows 1302 persons with Other White ethnicity including Polish (371), Portuguese (100), Romanian (129), Lithuanian (65), and Other Eastern European (65). Dr Philippa Brice, Associate Director of Research and Impact for Cambridgeshire and Peterborough Integrated Care Board said that the vaccination team had searched the archives but were “not aware of any documents or records that can shed light” on why uptake was below prediction. However, currently “for Primary Care research studies (not limited to vaccine studies), local practices are working to increase the availability of information in locally spoken languages” and this is helpful in increasing diverse uptake. (personal communication)

Thirty miles distant in Wisbech (Fenland) the Rosmini Centre supported Eastern European communities during the pandemic.[57] Staff and volunteers with the relevant languages attended centres, workplaces and multi-occupied housing to encourage vaccine uptake and testing. The Centre Manager Anita Grodkiewicz was not aware of any such initiative in Soham at the time.“COVID information was generally made available in other languages but unless you knew where to look you would not have been able to access it.” Amongst East Cambridgeshire locations, “Soham has the highest concentration of field and factory work undertaken by Central and Eastern Europeans. Social media websites specifically accessed by migrant workers at the time had misinformation resulting in vaccine resistance.” Poor transport links in rural East Cambridgeshire also meant that very few people travelled from Soham to the centre in Wisbech. (personal communication)

There are high proportions of Other White ethnicity in two Fenland MSOA: Wisbech North (25.4%) and Wisbech South & Peckover (21.3%), with respective uptake 0.63 0.67, fitted values 0.68 0.71 and `MixIS` predictions 0.67 0.71. In Boston there are three such MSOA: Fenside & Lister Way (41.6%), Boston Central & North (36.6%) and Skirbeck (22.5%), with respective uptake 0.49 0.45 0.67, fitted values 0.52 0.48 0.66 and `MixIS` predictions 0.51 0.48 0.65. Boston had a vaccination hub with translators.[58] Whilst uptake was below prediction in all but one of these MSOA, the discrepancies are much smaller than in Soham.

Soham is not exceptional when modelling the Booster / 3rd Injection (B3I) with uptake of the 2nd Primary as an additional smooth term. Other White ethnicity is now a positive predictor (Fig. 10 panel **h**, Fig. 9 panel **o**). Despite that shift the 2^nd^ Primary, closely aligned with the 1st Primary, dominates the prediction, lowering the uptake for MSOA with large Other White communities. By contrast, whilst Pakistani ethnicity was a mildly negative predictor for P1 (Fig. 4 panel **k**), it was sharply negative for B3I controlling for UP2 (Fig. 9 panel **l**).

Fratton West & Portsea (in Portsmouth) has znAged2024 = 2.1 and 44% of the population are students. For P1, this MSOA was most exceptional (Fig. 7). Its uptake 0.74 far exceeded the `MixIS` prediction 0.54 as well as the fitted value 0.59 which includes the positive South East and Portsmouth “random effects”, proximity to the Royal Navy and Royal Marines bases which together add 0.31 to the linear predictor. The Director of Public Health Helen Atkinson commented that this area with a large student population benefited from a pharmacy offering the vaccine 300m from the University of Portsmouth campus at convenient times for students. The Public Health team co-ordinated with University staff over a testing centre and vaccination clinics. (personal communication) The Director also used the local media to counter anti-vaccine arguments.[59] The Student Union produced a factsheet on vaccine myths in December 2020.[60] University staff conducted research into misinformation[61] and declining vaccine confidence[62]. When modelling B3I controlling for UP2, the smoother for znAged2024 rises sharply on the right (Fig. 9 panel **g**). Uptake shifted in favour of the Booster in areas with high student populations. In Fratton West & Portsea, uptake of B3I was slightly higher than the `MixIS` prediction.

Abbey Field (Colchester) was exceptional for P1, with uptake 0.87, fitted value 0.77 and `MixIS` prediction 0.77. Although proximity to an Army base was not a credible predictor overall, Abbey Field is adjacent to Merville Barracks which houses the elite Parachute Regiment, a possible explanation.

Leake & Butterwick (Boston) and Boughton & Selling (Swale) had respective uptakes 0.63 0.70, fitted values 0.76 0.83 and `MixIS` predictions 0.77 0.83. Both MSOA are coastal areas with high values of the population weighted road distance to a GP surgery, which may suggest reduced contact with the NHS. Sites distributing the primary vaccine were also distant. In Leake & Butterwick the nearest site was in Boston, 6.7 km from the MSOA centroid. In Boughton & Selling the nearest site was 5.2 km from the centroid.

Forest Row & Coleman’s Hatch (Wealden) has a high proportion of Level 4 (or higher) qualifications and low IMD. The East Sussex Director of Public Health Darrell Gale commented that the area had a pre-pandemic history of vaccine hesitancy “based on a range of views and beliefs, often related to more ‘natural health’ solutions to health problems”. (personal communication) The East Sussex County Council vaccination strategy[63] issued in March 2021 observed that “Forest Row & Coleman’s Hatch has significantly low uptake across all age groups. Forest Row has a known history of opposing views to vaccines and healthcare in general. Behavioural intervention and education may need to be focused in this area.” With `MixIS` prediction 0.83, uptake of the 1st Primary was 0.71. Uptake of B3I 0.56 nearly matched prediction 0.57, controlling for UP2.

In Salford Central & University, with uptake 0.66, fitted value 0.56 and `MixIS` prediction 0.55 there was evidence of student involvement in promoting vaccination [64] but it is unclear whether other unmodelled factors also contributed to the increased uptake.

The only clear outlier for B3I was Batley Carr & Mount Pleasant in Kirklees, with `MixIS` prediction 0.40 and uptake 0.31. This MSOA has an exceptionally large Indian community and a large Pakistani community. Including a univariate smoother for znIndian when modelling B3I makes little difference to the prediction for Batley Carr & Mount Pleasant, which remains exceptional.

This paper differs from the approach to modelling the booster in [10]. The uptake of P2 is included as a predictor for B3I, in order to consider whether apparent impacts on B3I may arise from earlier impacts on P1 and therefore P2. For example whilst [10] found “Other White” to be a negative predictor for B3I, the current approach shows that the negative response occurs with P1, and that when modelling B3I controlling for P2 the “Other White” response curve is positive. The models are now fully Bayesian, consider Local Authorities as nested within Regions and do not involve secondary modelling of the residuals. On the other hand, the Bayesian models use fewer predictors overall.

### Limitations

The choice of which covariates to include in ℳ1 represents a compromise. Extending the model slowed computation and sometimes caused divergences. ℳ1 and the various `MixIS` fits of models with a term or group deleted, all converged using the default priors. Other covariates in *dat1* might improve the fit and loo_pit_overlay. These could include population density, distance to the nearest GP surgery and likewise vaccination site - distinguishing between GP, pharmacy, hospital hub, vaccination centre - car ownership, unemployment, COVID-19 incidence before or during rollout, other ethnicities, industrial sector, other minor occupational groups (and the 18 included in ℳ1 and ℳ3 may not be optimal). However, whilst Fig. 1 shows that the indicator DV for density *<* 500/km^2^ is clearly linked to increased uptake, DV is strongly correlated (positively or negatively) with the ethnicities and age groups included in ℳ1. Adding DV to the model makes almost no difference. The visually strong apparent effect of lower population density is completely mediated through other model covariates.

Repeated sampling from a model is unlikely to yield identical results, unless all the `Stan`parameters are reset exactly. The results reported here differ slightly from those obtained when running the Additional files. For P1, when using `lookout` on a resample, the least extreme outlier Salford Central & University (with uptake above prediction) was replaced by the least extreme outlier Cannon Park & University (with uptake above prediction), an MSOA containing much of the University of Warwick which had extensive testing and vaccination on site. Other outliers were unchanged.

Describing an MSOA as a “possible outlier” only means that its uptake appeared unusually unlikely when a given sample was evaluated with particular leave-one-out techniques. The choice of probability threshold 0.01 in the `lookout` method applied to the matrix of mixic and `MixIS` predicted values is somewhat arbitrary, but it produced comparable output to the alternative method of fitting a GPD to the top 20% of mixic values themselves. However the limited aim was to identify MSOA with apparently unusual uptake deserving investigation, rather than to select a “best” criterion for identifying outliers. In those MSOA identified as exceptional for P1 other unmodelled issues may be involved including the availability of interpreters, the level of coordination between Public Health and Universities, the distance to GP surgeries and vaccination sites, the willingness of Public Health staff to tackle anti-vaccine arguments publicly, the “elite” status of certain military bases and pre-existing attitudes to vaccination.

The p-values reported in Table 2 are based on applying the technique used in loo but substituting the pointwise elpd_mixis values for elpd_loo. This seems plausible as `MixIS` is known to give more accurate elpd values in general.

Using the cumulative data as of 13 December 2023 meant the analysis could not consider the evolution of the uptake and its changing dependence on covariates. Although much more computation would be needed for the full archive, the geography could be restricted to a single region with cumulative data at selected times.

## 5 Conclusion

A Bayesian multilevel beta-binomial model, correcting for uncertainty in the Vaccine Registry, gives a good fit to the Middle Super Output Area data on COVID-19 vaccination uptake. Any success or failure to anticipate and address problems with distribution of the 1st Primary had lasting consequences, as area uptake of both the 2nd Primary and Booster / 3rd Injection were highly influenced by uptake of the 1st Primary. Vaccination planning for a future pandemic should highlight the initial phase. Differences among regional and local “random effects” are credible but should be evaluated in the context of demographic and socio-economic covariates, as the raw data can be misleading. Covariates associated with lower area uptake (of either 1st Primary or Booster / 3rd Injection) include the population proportions of international migrants, African & Other Black (combined), Caribbean, Pakistani, Roma, Romanian, and Other White. All of these groups need support to overcome the barriers highlighted in the COVID-19 Inquiry, including language and the Hostile Environment. In particular the Other White group, the ethnicity with the largest impact on area uptake, needs more active outreach including translation, and a network of NHS interpreters could help. The term “Vaccine hesitancy” is misleading, as it points to the patient instead of highlighting NHS and public health strategy and capacity to offer relevant provision for communities during the most critical period of vaccination, its outset.

## Data Availability

All data produced in the present work are contained in the manuscript and Supplement

## 6 Additional Files

Additional File 1: **dat1.txt** compressed data, use `load`(`“dat1.txt”,verbose=TRUE`)

Additional File 2: **dat1col** plain text list of dat1 column names and brief descriptions

Additional File 3: **nest.csv** csv table of levels of LTLA nested within REG

Additional File 4: **codeM1** plain text code for M1, M1mix and looM1

Additional File 5: **codeFigs1-5** plain text code for Figures 1, 2, 3, 4, 5

Additional File 6: **codeFig6** plain text code for Figure 6

Additional File 7: **codeFig7** plain text code for Figure 7

Additional File 8: **codeFig8** plain text code for Figure 8

Additional File 9: **codeM3** plain text code for M3, M3mix and looM3

Additional File 10: **codeFig9** plain text code for Figure 9

Additional File 11: **AddTab1.pdf** pdf file for Additional Table 1

Additional File 12: **AddFig1.pdf** pdf file for Additional Figure 1

Additional File 13: **AddFig2.pdf** pdf file for Additional Figure 2

Additional File 14: **AddFig3.pdf** pdf file for Additional Figure 3

Additional File 15: **codeAddFigs** plain text code for Additional Figures 1, 2, 3

Additional File 16: **codeFig10** plain text code for Figure 10

## 7 Abbreviations

*B3I*: Booster / 3rd Injection
*brms*: Bayesian Regression Modelling using ‘Stan’
*ebfmi*: Estimated Bayesian Fraction of Missing Information
*ecdf*: Empirical Cumulative Distribution Function
*elpd*: Expected Log Predictive Density
*EU*: European Union
*GAM*: Generalized Additive Model
*gam*: `mgcv` programme to fit Generalized Additive Models
*gamm4*: programme to fit Generalized Additive Mixed Models using ‘`mgcv`’ and ‘lme4’
*GPD*: Generalized Pareto Distribution
*hdi*: Highest Posterior Density Interval
*IMD*: Index of Multiple Deprivation
*loo*: Efficient Leave-One-Out Cross-Validation and WAIC for Bayesian Models
*LSHTM*: London School of Hygiene and Tropical Medicine
*LSOA*: Lower Super Output Area
*LTLA*: Lower Tier Local Authority
*MAD*: mean absolute deviation
*`mgcv`*: Mixed GAM Computation Vehicle
*MixIS*: Mixture Importance Sampling
*mixic*: mixture information criterion
*mixpred*: uptake predicted by MixIS
*MSOA*: Middle Super Output Area
*P1*: 1st Primary vaccination
*P2*: 2nd Primary vaccination
*R*: Environment for Statistical Computing
*R̂*: modified Gelman-Rubin statistic split-*R̂*
*R*^2^: Bayesian R-squared
*REG*: Census Region (England)
*SAGE*: UK Scientific Advisory Group for Emergencies
*Stan*: Software for Bayesian Data Analysis
*VR*: Vaccine Registry
*VRx*: Ratio of Vaccine Registry to Census population aged 12 and over

## 8 Declarations

### Ethics approval and consent to participate

Data analysed in this study is either publicly available or derived from public sources. I did not seek ethical approval for its use.

### Consent for publication

All individuals giving personal communications quoted in this study have consented to their publication.

### Availability of data and materials

All data generated or analysed during this study are included in this published article [and its supplementary information files].

### Competing interests

The authors declare that they have no competing interests.

### Funding

This work was unfunded.

### Authors’ contributions

GD designed the study, wrote the code, carried out the computations, drafted the manuscript and prepared the figures.

## Acknowledgements

Not applicable

## Notes

### Competing Interest Statement

The authors have declared no competing interest.

### Funding Statement

This study did not receive any funding

### Author Declarations

The study used ONLY openly available anonymous aggregated human data, available before the study. The UK COVID-19 Dashboard is now archived at https://ukhsa-dashboard.data.gov.uk/covid-19-archive-data-download. English indices of deprivation 2019 are available at https://www.gov.uk/government/statistics/english-indices-of-deprivation-2019. UK Census 2021 covariates are available at https://www.nomisweb.co.uk/query/select/getdatasetbytheme.asp?theme=93. Local authority data on the European Union (EU) Referendum (2016) is available from the Electoral Reform Commission https://www.electoralcommission.org.uk/research-reports-and-data/our-reports-and-data-past-elections-and-referendums/results-and-turnout-eu-referendum. Data on influenza vaccination uptake in persons over 65 is available at Unitary Authority level at https://assets.publishing.service.gov.uk/media/605c66b2d3bf7f2f0d94183e/LA_Seasonal_flu_vaccine_uptake_in_GP_patients-1_Sep_2020_to_28_Feb_2021.ods

### Summary of Updates

The treatment of the "random effects" for Region has been revised to centre each regional distribution by the distribution of the mean regional effect. Comparison of regions no longer involves permuting the difference in outputs. A similar revision is applied for nested local effects. Several figures in the manuscript and codes in the Supplementary material incorporate these revisions.

