## Additional files for "How COVID-19 vaccination in England varied by geography, demography, deprivation and registry: Bayesian ecological modelling": AddTab1.pdf

### Additional Table 1: Model $\mathcal{M}3$ parametric terms

| **term** | **text** | **mean** | **hdi** |
|---|---|---|---|
| znBREX | Brexit | -0.008 | (-0.012 , -0.004) |
| znX125 | MgrOthServ | -0.005 | (-0.007 , -0.002) |
| znX213 | ITProf | 0.007 | (0.004 , 0.010) |
| znX216 | ResearchDev | 0.005 | (0.003 , 0.006) |
| znX223 | NursMidwif | 0.003 | (0.001 , 0.004) |
| znX231 | TeachEduc | -0.003 | (-0.006 , -0.001) |
| znX242 | FinanceProf | 0.006 | (0.004 , 0.009) |
| znX331 | ProtectServ | 0.001 | (-0.001 , 0.003) |
| znX341 | ArtLitMedia | 0.003 | (0.000 , 0.005) |
| znX355 | SalesMarket | 0.005 | (0.002 , 0.007) |
| znX421 | Secretarial | -0.004 | (-0.006 , -0.003) |
| znX531 | ConstrBuild | -0.006 | (-0.008 , -0.004) |
| znX613 | CaringPers | -0.003 | (-0.006 , 0.000) |
| znX625 | B&BOwnProp | -0.001 | (-0.002 , 0.000) |
| znX711 | SalesAsst | -0.005 | (-0.008 , -0.003) |
| znX721 | CustomServ | -0.005 | (-0.007 , -0.003) |
| znX821 | RoadTprtDrv | -0.011 | (-0.014 , -0.008) |
| znX913 | ElemProcPlt | -0.001 | (-0.004 , 0.002) |
| znX925 | ElemStorag | -0.004 | (-0.006 , 0.000) |
| znLTC | LongTermCond | 0.010 | (0.008 , 0.013) |
| znFLU | FluVac65+ | 0.011 | (0.006 , 0.016) |
| Army | proximalArmy | -0.001 | (-0.010 , 0.008) |
| RAF | proximalRAF | -0.007 | (-0.018 , 0.004) |
| Navy | proximalNavy | 0.005 | (-0.011 , 0.020) |
| Marines | proximalMarine | -0.028 | (-0.049 , -0.008) |
| commDefence | communHousDefence | -0.374 | (-0.553 , -0.189) |
| commPrison | communHousPrison | 0.185 | (0.065 , 0.319) |
| znMigUK | internalUKmigrat | 0.007 | (0.005 , 0.009) |
| znNOPP | NoPassport | 0.029 | (0.025 , 0.034) |
