## Additional files for "How COVID-19 vaccination in England varied by geography, demography, deprivation and registry: Bayesian ecological modelling": codeM1

#code for M1, M1mix and looM1
#all required files are available from the mini-website
#results may vary slightly when refitting the models
#e.g. numerical values shown below are illustrative and will vary

#prepare the data needed to run the main model M1
#choose a working directory and navigate to it
#download dat1, dat1col, mcodx and mnamx into this directory
#dat1 is a dataframe with 6773 rows and 206 columns
#it contains all the data needed for M1, and much more
#dat1col lists the dat1 column names with brief descriptions
#dat1 is indexed by row names mcodx using MSOA geographic codes of the form "E0200xxxx"
#mnamx are the corresponding text names of the MSOA

#open R and load the packages needed for M1 and other computation

library(rstan)
rstan_options(auto_write = TRUE)
library(brms)
library(cmdstanr)
options(mc.cores=2, brms.normalize=TRUE,brms.backend="cmdstanr")
library(bayesplot)
library(posterior)
library(ggplot2)
library(gridExtra)
library(metR)
library(loo)
library(scales)
library(shinystan)
library("matrixStats")
library(mev)
library(lookout)
library(HDInterval)

#some helper functions

q025<-function(v) quantile(v,0.025)
q975<-function(v) quantile(v,0.975)
hdlo<-function(v) hdi(v,0.95)[1]
hdhi<-function(v) hdi(v,0.95)[2]
ee<-function(v) ecdf(v)(v)
nn<-function(v) qnorm(ee(v)-1/(2*length(v)))
zn<-function(v) as.numeric(scale(nn(v)))
h<-function(t) (1+exp(-t))^-1


#select and transform the data to be used in M1

#setwd("Desktop/phetest/brms/msoa/P1/fitt63.2/Supp")
load("dat1.txt",verbose=TRUE)

mcodx<-dat1$mcodx
mnamx<-dat1$mnamx
#MSOA geographic codes and names

P1<-dat1$P1
VR<-dat1$VR
REG<-dat1$REG
LTLA<-dat1$LTLA
znBREX<-zn(dat1$BREX)
znX125<-zn(dat1$X125)
znX213<-zn(dat1$X213)
znX216<-zn(dat1$X216)
znX223<-zn(dat1$X223)
znX231<-zn(dat1$X231)
znX242<-zn(dat1$X242)
znX331<-zn(dat1$X331)
znX341<-zn(dat1$X341)
znX355<-zn(dat1$X355)
znX421<-zn(dat1$X421)
znX531<-zn(dat1$X531)
znX613<-zn(dat1$X613)
znX625<-zn(dat1$X625)
znX711<-zn(dat1$X711)
znX721<-zn(dat1$X721)
znX821<-zn(dat1$X821)
znX913<-zn(dat1$X913)
znX925<-zn(dat1$X925)
znLTC<-zn(dat1$LTC)
znFLU<-zn(dat1$FLU)
Army<-dat1$Army
RAF<-dat1$RAF
Navy<-dat1$Navy
Marines<-dat1$Marines
commDefence<-dat1$commDefence
commPrison<-dat1$commPrison
znMigUK<-zn(dat1$MigUK)
znNOPP<-zn(dat1$NOPP)
znPROPFEM<-zn(dat1$PROPFEM)
znMigxUK<-zn(dat1$MigxUK)
znNoAnsRel<-zn(dat1$NoAnsRel)
znNoAnsSex<-zn(dat1$NoAnsSex)
znPROP1519<-zn(dat1$PROP_1519)
znPROP2024<-zn(dat1$PROP_2024)
znPROP55p<-zn(dat1$PROP_55p)
znLev01<-zn(dat1$NoQual+dat1$Lev1)
znAfrOB<-zn(dat1$AfrOB)
znCarib<-zn(dat1$Carib)
znPakistani<-zn(dat1$Pakistani)
znRoma<-zn(dat1$Roma)
znRomanian<-zn(dat1$Romanian)
znOthWh<-zn(dat1$OthWh)
znWhBr<-zn(dat1$WhBr)
znIMD<-zn(dat1$IMD)
znVRx<-zn(dat1$VRx)

#assemble datM1

datM1<-data.frame(P1,VR,REG,LTLA,znBREX,znX125,znX213,znX216,znX223,
znX231,znX242,znX331,znX341,znX355,znX421,znX531,
znX613,znX625,znX711,znX721,znX821,znX913,znX925,
znLTC,znFLU,Army,RAF,Navy,Marines,commDefence,commPrison,
znMigUK,znNOPP,znPROPFEM,znMigxUK,znNoAnsRel,znNoAnsSex,
znPROP1519,znPROP2024,znPROP55p,znLev01,znAfrOB,znCarib,
znPakistani,znRoma,znRomanian,znOthWh,znWhBr,znIMD,znVRx)

rownames(datM1)<-rownames(dat1)
#i.e. mcodx

dim(datM1)
sum(is.na(datM1))
#0  hopefully

#define the formula for M1
#LTLA nested within REG

formM1<-as.formula(P1 | vint(VR) ~ znBREX + znX125 + znX213 + znX216 + znX223 + znX231 + znX242 + znX331 + znX341 + znX355 + znX421 + znX531 + znX613 + znX625 + znX711 + znX721 + znX821 + znX913 + znX925 + znLTC + znFLU + Army + RAF + Navy + Marines + commDefence + commPrison + znMigUK + znNOPP + s(znPROPFEM, k = 7) + s(znMigxUK, k = 7) + s(znNoAnsRel, k = 7) + s(znNoAnsSex, k = 7) + s(znPROP1519, k = 7) + s(znPROP2024, k = 7) + s(znPROP55p, k = 7) + s(znLev01, k = 7) + s(znAfrOB, k = 7) + s(znCarib, k = 7) + s(znPakistani, k = 7) + s(znRoma, k = 7) + s(znRomanian, k = 7) + s(znOthWh, k = 7) + s(znWhBr, k = 7) + t2(znIMD, znVRx) + (1 | REG/LTLA) -1)

#convert to a brms formula with QR decomposition for the parametric terms

bfM1<-bf(formM1,decomp="QR")

#define the custom family and adaptation of stan code needed to interpret this
#as a beta_binomial model. see https://cran.r-project.org/web/packages/brms/vignettes/#brms_customfamilies.html#the-beta-binomial-distribution

beta_binomial2 <- custom_family(
  "beta_binomial2", dpars = c("mu", "phi"),
  links = c("logit", "log"),
  lb = c(0, 0), ub = c(1, NA),
  type = "int", vars = "vint1[n]"
)     

stan_funs <- "
  real beta_binomial2_lpmf(int y, real mu, real phi, int T) {
    return beta_binomial_lpmf(y | T, mu * phi, (1 - mu) * phi);
  }
  int beta_binomial2_rng(real mu, real phi, int T) {
    return beta_binomial_rng(T, mu * phi, (1 - mu) * phi);
  }
"

stanvars <- stanvar(scode = stan_funs, block = "functions")

posterior_epred_beta_binomial2 <- function(prep) {
  mu <- brms::get_dpar(prep, "mu")
  trials <- prep$data$vint1
  trials <- matrix(trials, nrow = nrow(mu), ncol = ncol(mu), byrow = TRUE)
  mu * trials
}

#specify M1 and fit it as below (may take 24 hrs on a 2-core 3GHz PC)

M1<-brm(bfM1,data=datM1,chains=4,family=beta_binomial2,stanvars=stanvars,
save_warmup=TRUE,iter=2000,warmup=1000,control=list(adapt_delta=0.8,max_treedepth=13))
saveRDS(M1,file="M1.RDS")

#uses default priors

#if resuming after fitting M1, closing and reopening the R session
#set the working directory, load the various programmes and run the next 3 lines
#M1<-readRDS("M1.RDS")
#datM1<-M1$data
#attach(datM1)

#DIAGNOSTICS
#see https://mc-stan.org/learn-stan/diagnostics-warnings.html
#and https://mc-stan.org/bayesplot/reference/index.html

#divergences
np<-nuts_params(M1)
lp<-log_posterior(M1)
mcmc_nuts_divergence(np,lp)

#rhat
draws<-as_draws_array(M1,inc_warmup=TRUE)
sam<-1001:2000
sumd<-summary(draws[sam,,])
range(sumd[,8])
ir<-which(sumd[,8]>1.01)
range(sumd[-ir,8])
sumd[ir,1]
unlist(sumd[ir,8])

#energy plot
mcmc_nuts_energy(np)

#E-BFMI
get_bfmi(M1$fit)
#measures the efficiency of Stan in exploring the energy levels in phase space
#E-BFMI < 0.3 would indicate a problem.

#posterior predictive density
vars<-variables(M1)
P1<-datM1$P1
VR<-datM1$VR
UP1<-P1/VR
pp<-matrix(NA,4000,6773)
phiv<-grep("phi",vars)

pospred<-posterior_epred(M1)
mumat<-pospred/outer(rep(1,4000),VR)
mat<-as.matrix(M1)
identical(vars,colnames(mat))

for (j in 1:4000)
{
muj<-mumat[j,]
phij<-as.numeric(mat[j,phiv])
aj<-phij*muj
bj<-phij*(1-muj)
pj<-rbeta(6773,aj,bj)
pp[j,]<-rbinom(6773,size=VR,prob=pj)
}

ppc_dens_overlay(P1,pp[1:100,])
denso<-ppc_dens_overlay(UP1,pp[1:100,]/outer(rep(1,100),VR))+
ggtitle("ppc_dens_overlay_UP1")
denso

#MAD
fit<-apply(mumat,2,mean)
MAD<-round(mean(abs(UP1-fit)),3)
MAD

#Bayesian R2
#see https://paulbuerkner.com/brms/reference/bayes_R2.brmsfit.html
#may take a minute to run
BR2<-bayes_R2(M1)
BR2<-round(BR2,3)

#parametric terms (Table 1)
#e.g. znBrex
vars[1]
mean(mat[,1])
hdi(mat[,1])

#"random effect" London vs North West

dif35<-mat[,phiv+3]-mat[,phiv+5]
mean(dif35)
hdi(dif35)


#"random effect" Allerdale vs Solihull

grep("Allerdale",vars)
grep("Solihull",vars)

difAS<-mat[,202]-mat[,350]
mean(difAS)
hdi(difAS)


#top 10 LTLA vs bottom 10 LTLA

matL<-mat[,phiv+9+1:307]
matLc<-matL-outer(rowMeans(matL),rep(1,307))

mLc<-colMeans(matLc)
oLc<-order(mLc)
colnames(matLc)[head(oLc,10)]
colnames(matLc)[tail(oLc,10)]

tab<-matrix(NA,10,10)
for (i in 1:10)
{
for (j in 1:10)
{
difij<-matLc[,oLc[308-i]]-matLc[,oLc[j]]
tab[i,j]<-hdi(difij)[1]
#lower limit of credible interval
}
}

tab
#all > 0

#contributions to interaction term for Harehills South and Ormskirk Town & East
#see codeFig6 for the function mterms based on decomp

mnamx[c(1620,1222)]
mterms(1620)[45]
mterms(1222)[45]


#MIXIS
#specify and run M1mix, to obtain the mixis elpd values and predicted values
#for background see https://mc-stan.org/loo/articles/loo2-mixis.html

cat(M1$model,file="M1.stan")
#save as M1mix.stan
#amend the model block with mixis code lines as below
#amend the generated quantities block to generate mu and log_lik as below
#save as M1mix.stan

#to amend the model block
#introduce a line declaring log_lik
#alter the section with the target loop
#leave the lines re priors unchanged

#the amended model block now reads (up to the lines re priors, omitted below):

model {
  // likelihood including constants
  if (!prior_only) {
    // initialize linear predictor term
    vector[N] log_lik = rep_vector(0.0, N);
    vector[N] mu = rep_vector(0.0, N);
    mu += XQ * bQ + Xs * bs + Zs_1_1 * s_1_1 + Zs_2_1 * s_2_1 + Zs_3_1 * s_3_1 + Zs_4_1 * s_4_1 + Zs_5_1 * s_5_1 + Zs_6_1 * s_6_1 + Zs_7_1 * s_7_1 + Zs_8_1 * s_8_1 + Zs_9_1 * s_9_1 + Zs_10_1 * s_10_1 + Zs_11_1 * s_11_1 + Zs_12_1 * s_12_1 + Zs_13_1 * s_13_1 + Zs_14_1 * s_14_1 + Zs_15_1 * s_15_1 + Zs_16_1 * s_16_1 + Zs_16_2 * s_16_2 + Zs_16_3 * s_16_3;
    for (n in 1:N) {
      // add more terms to the linear predictor
      mu[n] += r_1_1[J_1[n]] * Z_1_1[n] + r_2_1[J_2[n]] * Z_2_1[n];
    }
    mu = inv_logit(mu);
    for (n in 1:N) {
      log_lik[n] = beta_binomial2_lpmf(Y[n] | mu[n], phi, vint1[n]);
    }
    target += sum(log_lik);
    target += log_sum_exp(-log_lik);
  }
  // priors including constants
  

#to amend the generated quantities block
#copy the lines from the model block re log_lik and mu
#the amended generated quantities block now reads

generated quantities {
  // obtain the actual coefficients
  vector[K] b = XR_inv * bQ;
    vector[N] log_lik = rep_vector(0.0, N);
    vector[N] mu = rep_vector(0.0, N);
    mu += XQ * bQ + Xs * bs + Zs_1_1 * s_1_1 + Zs_2_1 * s_2_1 + Zs_3_1 * s_3_1 + Zs_4_1 * s_4_1 + Zs_5_1 * s_5_1 + Zs_6_1 * s_6_1 + Zs_7_1 * s_7_1 + Zs_8_1 * s_8_1 + Zs_9_1 * s_9_1 + Zs_10_1 * s_10_1 + Zs_11_1 * s_11_1 + Zs_12_1 * s_12_1 + Zs_13_1 * s_13_1 + Zs_14_1 * s_14_1 + Zs_15_1 * s_15_1 + Zs_16_1 * s_16_1 + Zs_16_2 * s_16_2 + Zs_16_3 * s_16_3;
    for (n in 1:N) {
      // add more terms to the linear predictor
      mu[n] += r_1_1[J_1[n]] * Z_1_1[n] + r_2_1[J_2[n]] * Z_2_1[n];
    }
    mu = inv_logit(mu);
    for (n in 1:N) {
      log_lik[n] = beta_binomial2_lpmf(Y[n] | mu[n], phi, vint1[n]);
    }
}


#make Cmdstanr model and run it (may take 24 hrs on 2-core 3GHz PC)

M1.mix<-cmdstan_model("M1mix.stan")
class(M1.mix)
M1.mix$print()

stand<-standata(M1)

mixM1<-M1.mix$sample(data=stand,chains=4,parallel_chains=2,refresh=100,
iter_warmup=1000,iter_sampling=1000,save_warmup=TRUE,max_treedepth=13,adapt_delta=0.9)
mixM1$save_object(file="mixM1.RDS")
#mixM1<-readRDS("mixM1.RDS")

#obtain elpd from mixis

draws<-mixM1$draws()
drawmat<-as_draws_matrix(draws)
dim(drawmat)

grep("log_lik",colnames(drawmat))[c(1,6773)]
dwLL<-drawmat[,924:7696]
colnames(dwLL)[c(1,6773)]

l_common_mix <- rowLogSumExps(-dwLL)
log_weights <- -dwLL - l_common_mix
elpd_mixis <- logSumExp(-l_common_mix) - rowLogSumExps(t(log_weights))
saveRDS(elpd_mixis,file="elpd_mixis_M1.RDS")
#elpd_mixis<-readRDS("elpd_mixis_M1.RDS")


#predict from mixis
lw<-log_weights
lse<-apply(lw,2,logSumExp)
lseo<-outer(rep(1,4000),lse)
lwn<-lw-lseo
w<-exp(lwn)
dim(w)
range(w)
#normalised weights
#in each column, these give the probabilities that a draw from the mixture resulted from #omitting the point corresponding to that column

dwmu<-drawmat[,7697:14469]
dim(dwmu)
colnames(dwmu)[c(1,6773)]
#mu values generated by mixis

emix<-apply(dwmu*w,2,sum)
#the predictions obtained using the normalised weights

#LOO
#see https://mc-stan.org/loo/

mat<-as.matrix(M1)
dim(mat)
identical(colnames(mat),variables(M1))
vars<-variables(M1)

pospred<-posterior_epred(M1)
pospredr<-pospred/outer(rep(1,4000),VR)
mumat<-pospredr
dim(mumat)
mnmumat<-apply(mumat,2,mean)

fit<-fitted(M1)[,1]/VR
range(fit-mnmumat)

phiv<-grep("phi",vars)

LL<-matrix(NA,4000,6773)
phimat<-drop(outer(mat[,phiv],rep(1,6773)))
alpha<-mumat*phimat
beta<-(1-mumat)*phimat

for (j in 1:6773)
{
LL[,j]<-lchoose(VR[j],P1[j])+lbeta(P1[j]+alpha[,j],VR[j]-P1[j]+beta[,j])-lbeta(alpha[,j],beta[,j])
}

eLL<-exp(LL)

cid<-c(rep(1,1000),rep(2,1000),rep(3,1000),rep(4,1000))
reff<-relative_eff(eLL,chain_id=cid)

looM1<-loo(LL,r_eff=reff,save_psis=TRUE)
saveRDS(looM1,file="looM1.RDS")
#readRDS("looM1.RDS")

looM1

looM1pt<-looM1$pointwise
elpd_loo<-looM1pt[,1]
ran<-range(c(elpd_mixis,elpd_loo))
plot(elpd_mixis,elpd_loo,xlim=ran,ylim=ran)
lines(ran,ran,col="red")

psis_object<-looM1$psis_object
log_ratios<--1*LL
eloo<-E_loo(mumat,psis_object,type="mean",log_ratios=log_ratios)
eloov<-eloo$value
save(emix,eloov,file="P1mixloov.RDS")
#load("P1mixloov.RDS",verbose=TRUE)
#see https://cran.r-project.org/web/packages/loo/refman/loo.html#E_loo

plot(emix,eloov,xlab="mixis_predict",ylab="loo_predict")
lines(range(emix),range(emix),col="red")

MADloo<-round(mean(abs(UP1-eloov)),3)
MADmix<-round(mean(abs(UP1-emix)),3)

ppc_loo_pit_overlay(P1,pp,samples=200,psis_object=looM1$psis)

#PIT is the Probability Integral Transform, approximated by ecdf
#if the observed values plausibly arose from the leave-one-out distribution
#then applying the PIT from the loo values to the observed values
#should give a uniform distribution
#the overlay tests this graphically

###
#model comparison with deleted term (Table 2)
#e.g. deleting (1|REG/LTLA)

#make Cmdstanr model and run it (may take 15 hrs on 2-core 3GHzPC)

M1<-readRDS("M1.RDS")
bfM1<-M1$formula
datM1<-M1$data

bfM1xRL<-update(bfM1,~.-(1|REG/LTLA)+1)
bfM1xRL<-bf(bfM1xRL,decomp="QR")

standxRL<-standata(bfM1xRL,datM1)
stanvars<-M1$stanvars
stancode(bfM1xRL,data=datM1,family="beta_binomial2",stanvars=stanvars,save_model="codexRL.stan")

#alter code and save as codexRLmix.stan

M1xRL.mix<-cmdstan_model("codexRLmix.stan")
class(M1xRL.mix)
M1xRL.mix$print()

mixM1xRL<-M1xRL.mix$sample(data=standxRL,chains=4,parallel_chains=2,refresh=100,
iter_warmup=1000,iter_sampling=1000,save_warmup=TRUE,
max_treedepth=13,adapt_delta=0.9)
mixM1xRL$save_object(file="mixM1xRL.RDS")
#mixM1xRL<-readRDS("mixM1xRL.RDS")

#14.7 hrs
#2 div
#maxtree 10

np<-nuts_params(mixM1xRL)
table(np$Value[np$Parameter=="divergent__"])
table(np$Value[np$Parameter=="treedepth__"])

draws<-mixM1xRL$draws()
dim(draws)
drawmat<-as_draws_matrix(draws)
dim(drawmat)

grep("log_lik",colnames(drawmat))[c(1,6773)]
dwLL<-drawmat[,292:7064]
dim(dwLL)
colnames(dwLL)[c(1,6773)]

l_common_mix <- rowLogSumExps(-dwLL)
log_weights <- -dwLL - l_common_mix
elpd_mixis_xRL <- logSumExp(-l_common_mix) - rowLogSumExps(t(log_weights))
saveRDS(elpd_mixis_xRL,file="elpd_mixis_xRL.RDS")
sum(elpd_mixis_xRL)
#-43045.03

elpd_mixis<-readRDS("elpd_mixis_M1.RDS")
sum(elpd_mixis)
#-41881.33

#as with loo_compare

#Delta
delta<-sum(elpd_mixis-elpd_mixis_xRL)
#1163.702

sddelta<-sd(elpd_mixis-elpd_mixis_xRL)*6773^0.5
#52.80415
#difference is highly significant
p=2*(1-pnorm(delta/sddelta))
scientific(p)

#dpar
delta/318
#3.659442

#how often does the full model fit better than the deleted model without (1|REG/LTLA)?
sum(elpd_mixis-elpd_mixis_xRL>0)/6773

#how often is the interaction term t2(znIMD,znVRx) the largest absolute term in the linear predictor?
#see codeFig6 for the function key10 based on mterms and decomp

nam<-c("znBREX","znX125","znX213","znX216","znX223",
"znX231","znX242","znX331","znX341","znX355","znX421","znX531","znX613",
"znX625","znX711","znX721","znX821","znX913","znX925","znLTC",
"znFLU","Army","RAF","Navy","Marines","commDefence","commPrison",
"znMigUK","znNOPP","znPROPFEM","znMigxUK","znNoAnsRel","znNoAnsSex","znPROP1519",
"znPROP2024","znPROP55p","znLev01","znAfrOB","znCarib",
"znPakistani","znRoma","znRomanian","znOthWh","znWhBr","znIMD:znVRx","REG","LTLA")


tabl<-matrix(0,10,47)
rownames(tabl)<-1:10
colnames(tabl)<-nam

pb<-txtProgressBar(min=1,max=6773,initial=1,style=3)
for (i in 1:6773)
{
setTxtProgressBar(pb,value=i)
ky<-colnames(key10(i))
mt<-match(ky,nam)
for (j in 1:10)
{
tabl[j,mt[j]]<-1+tabl[j,mt[j]]
}
}
write.table(tabl,file="keytabl")
#keytabl<-read.table("keytabl")
