## Additional files for "How COVID-19 vaccination in England varied by geography, demography, deprivation and registry: Bayesian ecological modelling": codeM3

#code for M3, M3mix and LOO
#all required files are available from the mini-website
#results may vary slightly when refitting the models

#prepare the data needed to run the main model M3
#choose a working directory and navigate to it
#download dat1, dat1col, mcodx and mnamx into this directory
#dat1 is a dataframe with 6773 rows and 206 columns
#it contains all the data needed for M3, and much more
#dat1col lists the dat1 column names with brief descriptions
#dat1 is indexed by row names mcodx using MSOA geographic codes of the form "E0200xxxx"
#mnamx are the corresponding text names of the MSOA

#open R and load the packages needed for M3 and other computation

library(rstan)
rstan_options(auto_write = TRUE)
library(brms)
library(cmdstanr)
options(mc.cores=2, brms.normalize=TRUE,brms.backend="cmdstanr")
library(bayesplot)
library(posterior)
library(ggplot2)
library(gridExtra)
library(metR)
library(loo)
library(scales)
library(shinystan)
library("matrixStats")
library(mev)
library(lookout)
library(HDInterval)

#some helper functions

q025<-function(v) quantile(v,0.025)
q975<-function(v) quantile(v,0.975)
hdlo<-function(v) hdi(v,0.95)[1]
hdhi<-function(v) hdi(v,0.95)[2]
ee<-function(v) ecdf(v)(v)
nn<-function(v) qnorm(ee(v)-1/(2*length(v)))
zn<-function(v) as.numeric(scale(nn(v)))
h<-function(t) (1+exp(-t))^-1


#select and transform the data to be used in M3

#setwd("Desktop/phetest/brms/msoa/P1/fitt63.2/Supp")
load("dat1.txt",verbose=TRUE)

mcodx<-dat1$mcodx
mnamx<-dat1$mnamx
#MSOA geographic codes and names

B3I<-dat1[,3]
P2<-dat1[,2]
VR<-dat1[,4]
UP2<-P2/VR

M1<-readRDS("M1.RDS")

bfM1<-M1$form
datM1<-M1$data

bfM3<-update(bfM1,B3I|vint(VR)~.+s(UP2,k=7))
bfM3<-bf(bfM3,decomp="QR")
datM3<-cbind(datM1,B3I,UP2)

stanvars<-M1$stanvars

#fit M3 (may take 24 hrs on 2-core 3GHz PC)

M3<-brm(bfM3,datM3,chains=4,family=beta_binomial2,stanvars=stanvars,
save_warmup=TRUE,iter=2000,warmup=1000,control=list(adapt_delta=0.8,max_treedepth=13))
saveRDS(M3,file="M3.RDS")
#M3<-readRDS("M3.RDS")

#if resuming after fitting M3, closing and reopening the R session
#set the working directory, load the various programmes and run the next 3 lines
#M3<-readRDS("M3.RDS")
#datM3<-M3$data
#attach(datM3)

#DIAGNOSTICS
#see https://mc-stan.org/learn-stan/diagnostics-warnings.html
#and https://mc-stan.org/bayesplot/reference/index.html

#divergences
np<-nuts_params(M3)
lp<-log_posterior(M3)
mcmc_nuts_divergence(np,lp)

#rhat
draws<-as_draws_array(M3,inc_warmup=TRUE)
sam<-1001:2000
sumd<-summary(draws[sam,,])
range(sumd[,8])
ir<-which(sumd[,8]>1.01)
range(sumd[-ir,8])
sumd[ir,1]
unlist(sumd[ir,8])

#energy plot
mcmc_nuts_energy(np)

#E-BFMI
get_bfmi(M3$fit)
#measures the efficiency of Stan in exploring the energy levels in phase space
#E-BFMI < 0.3 would indicate a problem.

#posterior predictive density
vars<-variables(M3)
UB3I<-B3I/VR
pp<-matrix(NA,4000,6773)
phiv<-grep("phi",vars)

posterior_epred_beta_binomial2 <- function(prep) {
  mu <- brms::get_dpar(prep, "mu")
  trials <- prep$data$vint1
  trials <- matrix(trials, nrow = nrow(mu), ncol = ncol(mu), byrow = TRUE)
  mu * trials
}

pospred<-posterior_epred(M3)
mumat<-pospred/outer(rep(1,4000),VR)
mat<-as.matrix(M3)
identical(vars,colnames(mat))

for (j in 1:4000)
{
muj<-mumat[j,]
phij<-as.numeric(mat[j,phiv])
aj<-phij*muj
bj<-phij*(1-muj)
pj<-rbeta(6773,aj,bj)
pp[j,]<-rbinom(6773,size=VR,prob=pj)
}

ppc_dens_overlay(B3I,pp[1:100,])
denso<-ppc_dens_overlay(UB3I,pp[1:100,]/outer(rep(1,100),VR))+
ggtitle("ppc_dens_overlay_UB3I")
denso

#MAD
fit<-apply(mumat,2,mean)
MAD<-round(mean(abs(UB3I-fit)),3)
MAD
#0.006

#Bayesian R2
#see https://paulbuerkner.com/brms/reference/bayes_R2.brmsfit.html
#may take a minute to run
BR2<-round(mean(bayes_R2(M3)[1,1]),3)
BR2
#0.997

#parametric terms
#e.g. znBrex
vars[1]
mean(mat[,1])
hdi(mat[,1])
#-0.008 (-0.012 , -0.004)

#London vs North West
dif35<-mat[,phiv+3]-mat[,phiv+5]
mean(dif35)
hdi(dif35)

#Rushmoor vs Blackburn with Darwen
matL<-mat[,phiv+9+1:307]
matLc<-matL-outer(rowMeans(matL),rep(1,307))
mLc<-colMeans(matLc)
oLc<-order(mLc)

vars[phiv+9+oLc[1]]
#Blackburn with Darwen
vars[phiv+9+oLc[307]]
#Rushmoor

difRB<-mat[,phiv+9+oLc[307]]-mat[,phiv+9+oLc[1]]
mean(difRB)
hdi(difRB)

#top 10 LTLA vs bottom 10 LTLA

tab<-matrix(NA,10,10)
for (i in 1:10)
{
for (j in 1:10)
{
difij<-matLc[,oLc[308-i]]-matLc[,oLc[j]]
tab[i,j]<-hdi(difij)[1]
#lower limit of credible interval
}
}

tab
#all > 0


#MIXIS
#specify and run M3mix, to obtain the mixis elpd values and predicted values
#for background see https://mc-stan.org/loo/articles/loo2-mixis.html

cat(M3$model,file="M3.stan")
#M3.stan is a text file
#save M3.stan as M3mix.stan
#amend the model block with mixis code lines as below
#amend the generated quantities block to generate mu and log_lik as below
#save as M3mix.stan

#to amend the model block
#introduce a line declaring log_lik
#alter the section with the target loop
#leave the lines re priors unchanged

#the amended model block now reads (up to the lines re priors, omitted below):

model {
  // likelihood including constants
  if (!prior_only) {
    // initialize linear predictor term
    vector[N] log_lik = rep_vector(0.0, N);
    vector[N] mu = rep_vector(0.0, N);
    mu += XQ * bQ + Xs * bs + Zs_1_1 * s_1_1 + Zs_2_1 * s_2_1 + Zs_3_1 * s_3_1 + Zs_4_1 * s_4_1 + Zs_5_1 * s_5_1 + Zs_6_1 * s_6_1 + Zs_7_1 * s_7_1 + Zs_8_1 * s_8_1 + Zs_9_1 * s_9_1 + Zs_10_1 * s_10_1 + Zs_11_1 * s_11_1 + Zs_12_1 * s_12_1 + Zs_13_1 * s_13_1 + Zs_14_1 * s_14_1 + Zs_15_1 * s_15_1 + Zs_16_1 * s_16_1 + Zs_16_2 * s_16_2 + Zs_16_3 * s_16_3 + Zs_17_1 * s_17_1;
    for (n in 1:N) {
      // add more terms to the linear predictor
      mu[n] += r_1_1[J_1[n]] * Z_1_1[n] + r_2_1[J_2[n]] * Z_2_1[n];
    }
    mu = inv_logit(mu);
    for (n in 1:N) {
      log_lik[n] = beta_binomial2_lpmf(Y[n] | mu[n], phi, vint1[n]);
    }
    target += sum(log_lik);
    target += log_sum_exp(-log_lik);
  }
  // priors including constants
  

#to amend the generated quantities block
#copy the lines from the model block re log_lik and mu
#the amended generated quantities block now reads

generated quantities {
  // obtain the actual coefficients
  vector[K] b = XR_inv * bQ;
    vector[N] log_lik = rep_vector(0.0, N);
    vector[N] mu = rep_vector(0.0, N);
    mu += XQ * bQ + Xs * bs + Zs_1_1 * s_1_1 + Zs_2_1 * s_2_1 + Zs_3_1 * s_3_1 + Zs_4_1 * s_4_1 + Zs_5_1 * s_5_1 + Zs_6_1 * s_6_1 + Zs_7_1 * s_7_1 + Zs_8_1 * s_8_1 + Zs_9_1 * s_9_1 + Zs_10_1 * s_10_1 + Zs_11_1 * s_11_1 + Zs_12_1 * s_12_1 + Zs_13_1 * s_13_1 + Zs_14_1 * s_14_1 + Zs_15_1 * s_15_1 + Zs_16_1 * s_16_1 + Zs_16_2 * s_16_2 + Zs_16_3 * s_16_3 + Zs_17_1 * s_17_1;
    for (n in 1:N) {
      // add more terms to the linear predictor
      mu[n] += r_1_1[J_1[n]] * Z_1_1[n] + r_2_1[J_2[n]] * Z_2_1[n];
    }
    mu = inv_logit(mu);
    for (n in 1:N) {
      log_lik[n] = beta_binomial2_lpmf(Y[n] | mu[n], phi, vint1[n]);
    }
}

#make Cmdstanr model and run it (may take 30 hrs on 2-core 3GHz PC)

M3.mix<-cmdstan_model("M3mix.stan")
class(M3.mix)
M3.mix$print()

stand<-standata(M3)

mixM3<-M3.mix$sample(data=stand,chains=4,parallel_chains=2,refresh=100,
iter_warmup=1000,iter_sampling=1000,save_warmup=TRUE,max_treedepth=13,adapt_delta=0.9)
mixM3$save_object(file="mixM3.RDS")
#mixM3<-readRDS("mixM3.RDS")

#obtain elpd from mixis

draws<-mixM3$draws()
drawmat<-as_draws_matrix(draws)
dim(drawmat)

grep("log_lik",colnames(drawmat))[c(1,6773)]
dwLL<-drawmat[,936:7708]
colnames(dwLL)[c(1,6773)]

l_common_mix <- rowLogSumExps(-dwLL)
log_weights <- -dwLL - l_common_mix
elpd_mixis <- logSumExp(-l_common_mix) - rowLogSumExps(t(log_weights))
saveRDS(elpd_mixis,file="elpd_mixis_M3.RDS")
#elpd_mixis<-readRDS("elpd_mixis_M3.RDS")

#predict from mixis
lw<-log_weights
lse<-apply(lw,2,logSumExp)
lseo<-outer(rep(1,4000),lse)
lwn<-lw-lseo
w<-exp(lwn)
dim(w)
range(w)
#normalised weights
#in each column, these give the probabilities that a draw from the mixture resulted from #omitting the point corresponding to that column

dwmu<-drawmat[,7709:14481]
dim(dwmu)
colnames(dwmu)[c(1,6773)]
#mu values generated by mixis

emix<-apply(dwmu*w,2,sum)
#the predictions obtained using the normalised weights

#LOO
#see https://mc-stan.org/loo/

mat<-as.matrix(M3)
dim(mat)
identical(colnames(mat),variables(M3))
vars<-variables(M3)

pospred<-posterior_epred(M3)
pospredr<-pospred/outer(rep(1,4000),VR)
mumat<-pospredr
dim(mumat)
mnmumat<-apply(mumat,2,mean)

fit<-fitted(M3)[,1]/VR
range(fit-mnmumat)

phiv<-grep("phi",vars)

LL<-matrix(NA,4000,6773)
phimat<-drop(outer(mat[,phiv],rep(1,6773)))
alpha<-mumat*phimat
beta<-(1-mumat)*phimat

for (j in 1:6773)
{
LL[,j]<-lchoose(VR[j],B3I[j])+lbeta(B3I[j]+alpha[,j],VR[j]-B3I[j]+beta[,j])-lbeta(alpha[,j],beta[,j])
}

eLL<-exp(LL)

cid<-c(rep(1,1000),rep(2,1000),rep(3,1000),rep(4,1000))
reff<-relative_eff(eLL,chain_id=cid)

looM3<-loo(LL,r_eff=reff,save_psis=TRUE)
saveRDS(looM3,file="looM3.RDS")
#looM3<-readRDS("looM3.RDS")

looM3

looM3pt<-looM3$pointwise
elpd_loo<-looM3pt[,1]
ran<-range(c(elpd_mixis,elpd_loo))
plot(elpd_mixis,elpd_loo,xlim=ran,ylim=ran)
lines(ran,ran,col="red")

psis_object<-looM3$psis_object
log_ratios<--1*LL
eloo<-E_loo(mumat,psis_object,type="mean",log_ratios=log_ratios)
eloov<-eloo$value
#see https://cran.r-project.org/web/packages/loo/refman/loo.html#E_loo

save(emix,eloov,file="B3Imixloov.RDS")
#load("B3Imixloov.RDS",verbose=TRUE)

plot(emix,eloov,xlab="mixis_predict",ylab="loo_predict")
lines(range(emix),range(emix),col="red")

MADloo<-round(mean(abs(UB3I-eloov)),3)
MADmix<-round(mean(abs(UB3I-emix)),3)

MADloo
MADmix

ppc_loo_pit_overlay(B3I,pp,samples=200,psis_object=looM3$psis)

#PIT is the Probability Integral Transform, approximated by ecdf
#if the observed values plausibly arose from the leave-one-out distribution
#then applying the PIT from the loo values to the observed values
#should give a uniform distribution
#the overlay tests this graphically

cs<-conditional_smooths(M3)
saveRDS(cs,file="csM3.RDS")
#cs<-readRDS("csM3.RDS")

###
#adding znIndian

znIndian<-zn(dat1$Indian)

bfM3<-M3$formula

bfM3I<-update(bfM3,~.+s(znIndian,k=7))
bfM3I<-bf(bfM3I,decomp="QR")
datM3I<-cbind(datM3,znIndian)

stanvars<-M3$stanvars

#fit M3I (may take 28 hrs on 2-core 3GHz PC)

M3I<-brm(bfM3I,datM3I,chains=4,family=beta_binomial2,stanvars=stanvars,
save_warmup=FALSE,iter=2000,warmup=1000,control=list(adapt_delta=0.8,max_treedepth=13))
saveRDS(M3I,file="M3I.RDS")
#M3I<-readRDS("M3I.RDS")

#28.4 hrs
#2 div
#max treedepth 13

np<-nuts_params(M3)
lp<-log_posterior(M3)
mcmc_nuts_divergence(np,lp)

UB3I<-B3I/VR

posterior_epred_beta_binomial2 <- function(prep) {
  mu <- brms::get_dpar(prep, "mu")
  trials <- prep$data$vint1
  trials <- matrix(trials, nrow = nrow(mu), ncol = ncol(mu), byrow = TRUE)
  mu * trials
}

fit<-fitted(M3I)[,1]/VR
MAD<-round(mean(abs(UB3I-fit)),3)
BR2<-round(bayes_R2(M3I)[,1],3)

plot(fit,UB3I,main="M3I",col=rgb(0,0,1,0.3))
points(fit[1525],UB3I[1525],pch=16,col=2)
text(fit[1525],UB3I[1525],pos=4,label=mnamx[1525])
text(0.2,0.8,pos=4,label="MAD")
text(0.28,0.8,pos=4,label=MAD)
text(0.2,0.75,pos=4,label="BR2")
text(0.28,0.75,pos=4,label=BR2)
