## Additional files for "How COVID-19 vaccination in England varied by geography, demography, deprivation and registry: Bayesian ecological modelling": dat1col

dat1 columns
brief definitions

1   P1: number of people who received the 1st Primary
2   P2: number of people who received the 2nd Primary
3   B3I: number of people who received the Booster / Third Injection
4   VR: vaccine registry
5   VRx: ratio VR / census population aged 12 and over
6   ethtot: total population shown in Census table ts021
7   REG: Region
8   LTLA: Lower Tier Local Authority
9   IMD: Index of Multiple Deprivation
10  IMDG: Deprivation Index - Barriers to Housing and Services
11  BARGP: Mean distance to GP surgery (km)
12  DEN: Population density per km2
13  DV: Indicator for DEN < 500/km2 (TRUE = 2, FALSE = 1)
14  PROPFEM: Proportion Female
15  PROP_1519: prop Aged 15-19
16  PROP_2024: prop Aged 20-24
17  PROP_2534: prop Aged 25-34
18  PROP_3554: prop Aged 35-54
19  PROP_55p: prop Aged 55 and over
20  PROP_4044: prop Aged 40-44
21  PROP_4549: prop Aged 45-49
22  PROP_5054: prop Aged 50-54
23  PROP_5559: prop Aged 55-59
24  PROP_6064: prop Aged 60-64
25  PROP_6569: prop Aged 65-69
26  PROP_7074: prop Aged 70-74
27  PROP_7579: prop Aged 75-79
28  PROP_8084: prop Aged 80-84
29  PROP_85p: prop Aged 85 and over
30  WhBr: prop White British ethnicity
31  Bangladeshi: prop Bangladeshi ethnicity
32  Chinese: prop Chinese ethnicity
33  Indian: prop Indian ethnicity
34  Pakistani: prop Pakistani ethnicity
35  OthAs: prop Other Asian ethnicity
36  AfrOB: prop Black African or Other Black ethnicity (combined)
37  Carib: prop Black Caribbean ethnicity
38  WhAs: prop Mixed White and Asian ethnicity
39  WhBlAfr: prop Mixed White and Black African ethnicity
40  WhBlCar: prop Mixed White and Black Caribbean ethnicity
41  OthMx: prop Other Mixed ethnicity
42  WhIri: prop White Irish ethnicity
43  GypIrT: prop Gypsy or Irish Traveller ethnicty
44  Roma: prop Roma ethnicity
45  OthWh: prop Other White ethnicity
46  Arab: prop Arab ethnicity
47  Other: prop Other ethnicty
48  Filipino: prop Filipino ethnicty
49  Nigerian: prop Nigerian ethnicity
50  Polish: prop Polish ethnicity
51  Romanian: prop Romanian ethnicity
52  Portuguese: prop Portuguese ethnicity
53  Spanish: prop Spanish ethnicity
54  EuroMix: prop European Mixed ethnicity
55  Bulgarian: prop Bulgarian ethnicity
56  Latvian: prop Latvian ethnicity
57  Lithuanian: prop Lithuanian ethnicity
58  NoRel: prop No Religion
59  Christian: prop Christian religion
60  Buddhist: prop Buddhist religion
61  Hindu: prop Hindu religion
62  Jewish: prop Jewish religion
63  Muslim: prop Muslim religion
64  Sikh: prop Sikh religion
65  OthRel: prop Other religion
66  NoAnsRel: prop No Answer to census question on religion
67  PROPA: Employed proportion in Agriculture, Forestry and fishing
68  PROPB: Emp prop in Mining and Quarrying
69  PROPC: Emp prop in Manufacturing
70  PROPD: Emp prop in Electricity, gas, steam and air conditioning supply
71  PROPE: Emp prop in Water supply; Sewerage, Waste management and Remediation
72  PROPF: Emp prop in Construction
73  PROPG: Emp prop in Wholesale and retail trade
74  PROPH: Emp prop in Transport and Storage
75  PROPI: Emp prop in Accommodation and Food Service
76  PROPJ: Emp prop in Information and Communication
77  PROPK: Emp prop in Finance and Insurance
78  PROPL: Emp prop in Real Estate
79  PROPM: Emp prop in Professional, Scientific and Technical
80  PROPN: Emp prop in Admin and Support
81  PROPO: Emp prop in Public administration and defence
82  PROPP: Emp prop in Education
83  PROPQ: Emp prop in Human Health and Social Work
84  Mgr: Emp prop Managers, directors and senior officials
85  Prof: Emp prop Professional
86  AssocProf: Emp prop Assoc professional and technical
87  AdminSec: Emp prop Administrative and Secretarial
88  Skilled: Emp prop skilled trades
89  CareLeis: Emp prop Caring, leisure and other service
90  Sales: Emp prop Sales and customer services
91  Process: Emp prop Process, pland and machine operatives
92  Element: Emp prop Elementary occupations
93  X112: Emp prop ProdMgr
94  X113: Emp prop FunctMgr
95  X115: Emp prop RetailWholMgr
96  X116: Emp prop SnrProtServ
97  X117: Emp prop HlthSocWkMgr
98  X121: Emp prop AgricMgr
99  X125: Emp prop MgrOthServ
100 X211: Emp prop NatSocSciProf
101 X212: Emp prop EnginProf
102 X213: Emp prop ITProf
103 X215: Emp prop ConservEnvirProf
104 X216: Emp prop ResearchDev
105 X222: Emp prop Therapy
106 X223: Emp prop NursMidwif
107 X224: Emp prop Vets
108 X225: Emp prop OthHlthProf
109 X231: Emp prop TeachEduc
110 X242: Emp prop FinanceProf
111 X246: Emp prop WelfareProf
112 X247: Emp prop Librarian
113 X249: Emp prop MediaProf
114 X311: Emp prop SciEngTecn
115 X312: Emp prop CADArchTec
116 X322: Emp prop WelfarAPro
117 X324: Emp prop VetNurs
118 X331: Emp prop ProtServ
119 X341: Emp prop ArtLitMedia
120 X342: Emp prop Design
121 X343: Emp prop SportsFit
122 X351: Emp prop TportAPro
123 X352: Emp prop LegalAPro
124 X355: Emp prop SalesMarket
125 X356: Emp prop PublSerAPro
126 X412: Emp prop AdminFinan
127 X415: Emp prop OthAdmin
128 X421: Emp prop Secretarial
129 X511: Emp prop Agric
130 X523: Emp prop VehicTrades
131 X524: Emp prop ElectrTrades
132 X525: Emp prop SkillMetElec
133 X531: Emp prop ConstrBuild
134 X532: Emp prop BuildFinish
135 X544: Emp prop OthSkilled
136 X612: Emp prop AnimalCare
137 X613: Emp prop CaringPers
138 X625: Emp prop B&BOwnProp
139 X711: Emp prop SalesAsst
140 X712: Emp prop SalesRelat
141 X713: Emp prop Shopkeepr
142 X721: Emp prop CustomServ
143 X813: Emp prop PlantMachOp
144 X821: Emp prop RoadTprtDrv
145 X911: Emp prop ElemAgric
146 X913: Emp prop ElemProcPlt
147 X921: Emp prop ElemAdmin
148 X923: Emp prop ElemSecur
149 X925: Emp prop ElemStorag
150 X926: Emp prop OthElemServ
151 NoQual: prop No qualification
152 Lev1: prop Level 1
153 Lev2: prop Level 2
154 Appren: prop Apprentice
155 Lev3: prop Level 3
156 Lev4: prop Level 4
157 OthQual: prop Other qualification
158 Straight: prop Straight
159 GayLes: prop Gay / Lesbian
160 Bi: prop Bisexual
161 OthSexOr: prop Oth Sexual Orientation
162 NoAnsSex: prop No Answer to census question on sexuality
163 DIS1: prop Disabled under the Equality Act - limited a lot
164 DIS2: prop Disabled under the Equality Act - limited a little
165 LTC: prop Not Disabled under Equality Act - Long Term Condition
166 FLU: prop Aged 65+ with flu vaccination (from LTLA data)
167 INC1: mean incidence rate 1 Oct 2020 - 31 Dec 2020
168 INC2: mean incidence rate 1 Jan 2021 - 31 Mar 2021
169 INC3: mean incidence rate 1 Apr 2021 - 30 Jun 2021
170 commDefence: prop housed in communal Defence establishment
171 commPrison: prop housed in communal Prison
172 commEduc: prop housed in communal Educational establishment
173 PHP: Public Health allocation 2020-21 per Vaccine Registry (Upper Tier Loc Auth) 
174 PHX: percentage change in Public Health allocation from 2019-2020 to 2020-21
175 po8: prop of households with 8 or more people
176 TENU: prop of households owned or shared ownership
177 HHND: prop of households not deprived in any dimension
178 HH1: prop households deprived in one dimension
179 HH2: prop households deprived in two dimensions
180 HH3: prop households deprived in three dimensions
181 HH4: prop households deprived in four dimensions
182 ONEP: prop one peron households
183 ONEP66: prop one peron households aged 66+
184 ONEPOTH: prop one peron households aged <66
185 ACTIV: prop aged 16+ and economically active (incl full-time students)
186 EMP: prop aged 16+ econ active and in employment (incl full-time students)
187 UNEM: prop aged 16+ econ active and unemployed (incl full-time students)
188 INAC: prop aged 16+ econ inactive
189 STUDG: prop of those aged 12+ who are students
190 NVWK: prop aged 16+ not currently in employment who have never worked
191 MigUK: prop migrant within UK in last 12 months
192 MigxUK: prop migrant from outside UK in last 12 months
193 NOPP: prop with no passport
194 LEP: prop with Limited English Proficiency
195 NOENGMN: prop households where no-one has English as main language
196 AFVET: prop previously served in UK armed forces
197 DisGP: shortest distance from centroid to GP dispensing vaccine Apr 2021
198 DisPHM: shortest distance from centroid to pharmacy dispensing vaccine Apr 2021
199 DisHH: shortest distance from centroid to hospital hub dispensing vaccine Apr 2021
200 DisVXC: shortest distance from centroid to vaccination centre Apr 2021
201 NoCar: proportion of households with no car or van
202 Army: indicator for proximity to British Army base
203 RAF: indicator for proximity to RAF base
204 Navy: indicator for proximity to Royal Navy base
205 Marines: indicator for proximity to Royal Marines base
206 BREX: proportion voting Leave in EU Referendum (projected from LTLA value)
207 mnamx: MSOA name
208 mcodx: MSOA code
