## Supplementary figures and images for "How COVID-19 vaccination in England varied by geography, demography, deprivation and registry: Bayesian ecological modelling"

### AddFig1.pdf

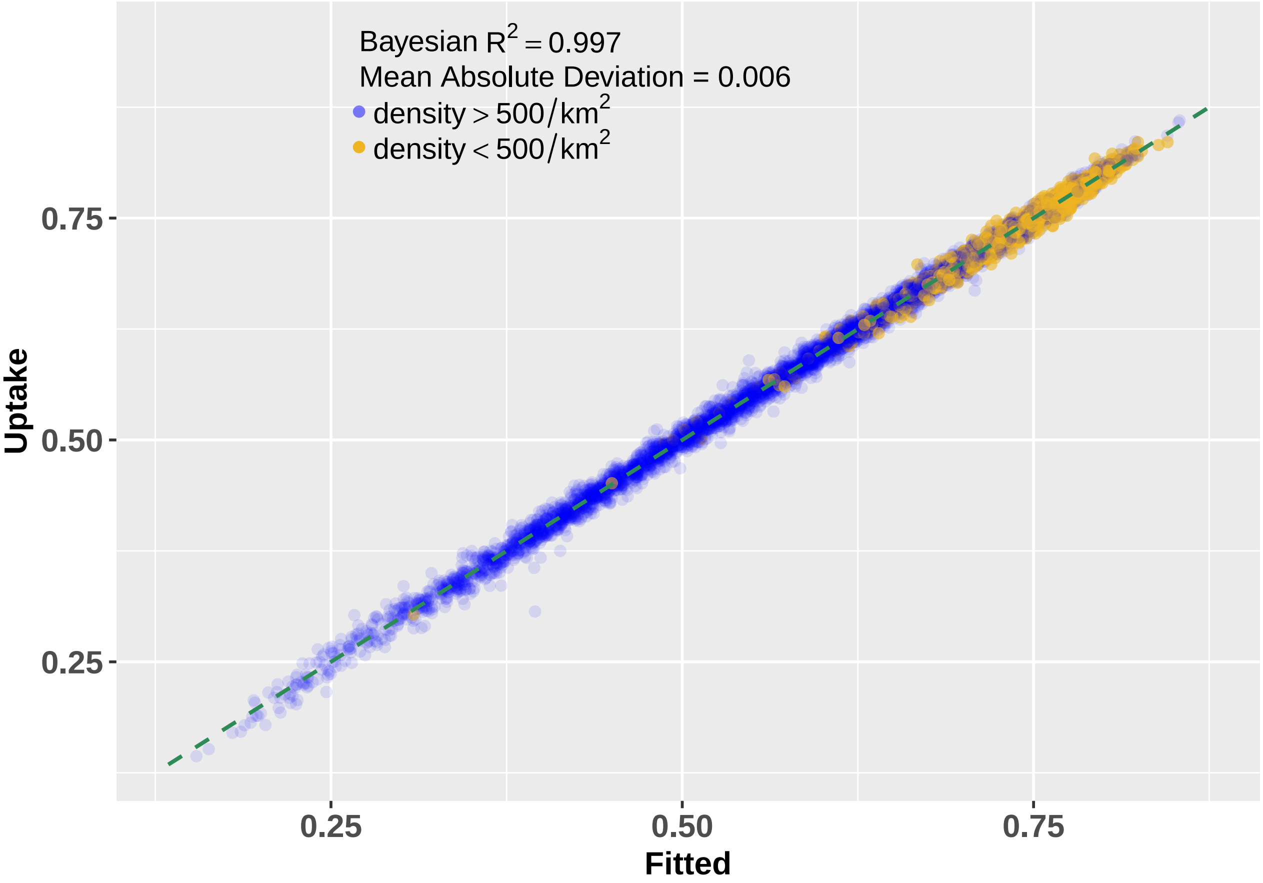

### AddFig2.pdf

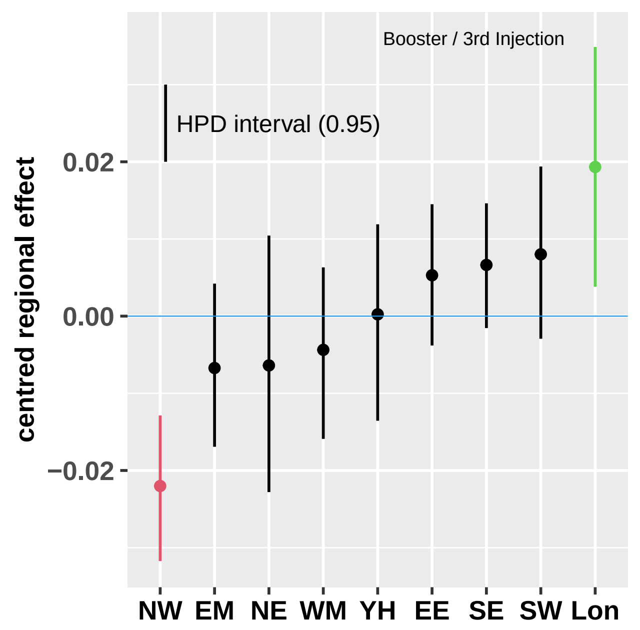

### AddFig3.pdf

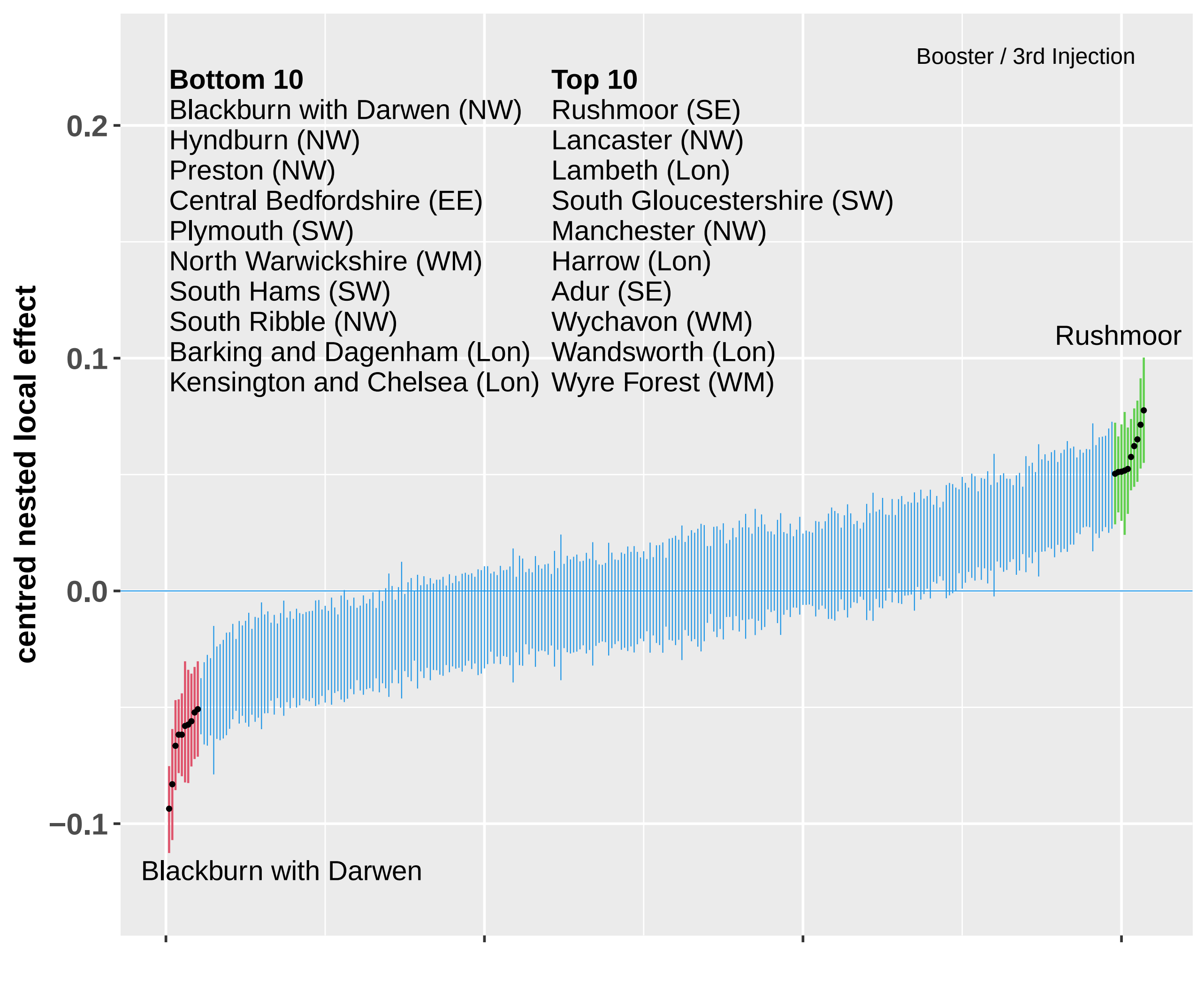
